# Depression increases both glycated haemoglobin testing frequency and risk of all-cause mortality in type 2 diabetes

**DOI:** 10.1101/2025.10.06.25337242

**Authors:** Dale Handley, Renu Bala, Katie G Young, Chris Wai Hang Lo, Jessica Tyrrell, Cathryn M Lewis, Alexandra C Gillett

**Affiliations:** Social, Genetic and Developmental Psychiatry Centre, Institute of Psychiatry, Psychology and Neuroscience, King’s College London, London, UK; Clinical and Biomedical Science, Institute of Health and Life Sciences, University of Exeter, Exeter, UK; NIHR Maudsley Biomedical Research Centre, South London and Maudsley NHS Trust, London, UK; Department of Medical and Molecular Genetics, Faculty of Life Sciences and Medicine, King’s College London, London, UK

## Abstract

**Background:** Frequent and consistent Glycated Haemoglobin (HbA1c) measurement is required for effective management of Type 2 diabetes (T2D). Co-morbid depression is an important driver of poor health and mortality in individuals with T2D. Little is known about how engagement with primary care for T2D management differs in individuals with co-morbid depression, and how this impacts all-cause mortality.

**Methods:** Adults with T2D were identified in the Clinical Practice Research Datalink Aurum dataset. Glycaemic testing frequency was defined as the number of HbA1c measurements per year after diagnosis. Conway-Maxwell-Poisson mixed effects models were used to test the associations between glycaemic testing frequency and T2D co-morbidities, pre-existing and incident depression (after T2D diagnosis), and depression severity. Cox proportional hazard models were used to determine the association of these variables with all-cause mortality.

**Results:** In total, 470,225 individuals with T2D were included. Of these, 20.1% and 6.1% had pre-existing and incident depression, respectively. Younger age, male sex, and non-White self-reported ethnicity were robustly associated with lower glycaemic testing frequency. In contrast, higher deprivation, pre-existing depression (IRR:1.06, CI: 1.05 – 1.07, p = 6.5×10-11), incident depression (IRR:1.04, CI: 1.03 – 1.05, p = 5.5×10-11) and a higher incident comorbidity score were associated with more frequent HbA1c testing. Pre-existing depression, incident depression, and depression severity were associated with increased risk of all-cause mortality after adjusting for HbA1c testing frequency (incident depression HR: 1.41, CI: 1.37 – 1.45, p = 3.7×10-97). In the entire cohort, higher HbA1c testing frequency was associated with reduced risk of all-cause mortality (HR: 0.93, 95% CI: 0.92 – 0.94, p = 3.60×10-82).

**Conclusion:** Individuals with T2D and depression have more frequent glycaemic testing yet still face a higher risk of mortality. This suggests that increased engagement with primary care for glycaemic monitoring alone is not sufficient to offset the broader health risks associated with depression. Targeted interventions beyond increased routine diabetes monitoring may be necessary to reduce excess mortality among individuals managing both T2D and depression.

## Introduction

Type 2 diabetes mellitus (T2D) is a chronic and progressive metabolic disorder that affects over 500 million people globally, a number that has increased by more than 90.5% since 1990 (1). It is a leading contributor to global morbidity and mortality, primarily through its associations with cardiovascular disease, renal failure, and other complications. T2D is characterised by insulin resistance and impaired insulin secretion, leading to chronically elevated blood glucose levels. However, the risk of adverse outcomes can be mitigated by improving glycaemic control, making it a fundamental part of effective T2D management.

Glycated Haemoglobin (HbA1c) measures average blood glucose levels over the past two to three months and is a key biomarker for glycaemic management. In the United Kingdom (UK), National Institute for Health and Care Excellence (NICE) guidelines recommend measuring HbA1c in primary care every three to six months to achieve optimal glycaemic control, with measurement frequency guided by previous HbA1c levels (2). Achieving and maintaining glycaemic control through frequent HbA1c testing has been shown to substantially reduce the risk of both vascular complications and mortality (3–5). Despite its clear importance in T2D management, it remains unclear whether psychiatric conditions, such as depression, impact engagement with HbA1c testing in primary care.

There is a complex bidirectional relationship between T2D and depression, with depression diagnosed prior to T2D diagnosis (i.e., pre-existing depression) and after T2D diagnosis (i.e., incident depression) playing important roles in T2D disease aetiology and progression (6). Pre-existing depression increases the risk of developing T2D through multiple mechanisms, including associations with smoking and substance use, unhealthy diet, and physical inactivity, which contribute to poor metabolic health (7). In addition, common comorbidities of depression, such as obesity and insomnia, are associated with increased risk of developing T2D (8). There are also shared biochemical processes, such as chronic low-grade inflammation and dysregulation of the hypothalamic-pituitary-adrenal (HPA) axis, which impair insulin sensitivity and glucose regulation and promote visceral fat accumulation (9).

Conversely, individuals with T2D are at increased risk of developing depression, although the mechanisms are less well understood. Diabetes-related complications, such as neuropathy or stroke, can result in chronic pain and reduced physical functioning, lowering quality of life and potentially contributing to the onset of depression (8). The requirements of managing and living with T2D, including dietary restrictions, adherence to medication regimens, and fear of developing complications, can cause substantial emotional distress (11). Finally, chronic hyperglycaemia and the subsequent associated inflammatory processes may contribute to cognitive decline and mood dysregulation, further increasing risk of developing depression (12).

Individuals with co-morbid depression and T2D have poorer glycaemic control and consequently, higher risks of vascular complications, premature mortality, and suicide (13). However, the mechanisms through which depression worsens T2D prognosis are understudied. While it is established that individuals with co-morbid depression and diabetes have worse self-management of diabetes, including lower medication adherence, another possibility is that individuals with these conditions attend HbA1c testing appointments less frequently than those without depression (14,15). Reduced testing could lead to suboptimal primary care management, resulting in poorer glycaemic control and a higher risk of complications and mortality.

Although individuals with depression typically have higher levels of overall healthcare utilisation, likely due to poorer health, most studies to date have focused on treatment adherence and self-management (16). To our knowledge, no studies which have examined the effect of depression on primary care-based glycaemic testing, and how this influences the risk of mortality in individuals with T2D.

To address this gap, we used the Clinical Practice Research Datalink (CPRD) Aurum dataset, which contains records from over 50 million active, transferred, and deceased patients. Specifically, we investigated whether pre-existing and incident depression in individuals with T2D is associated with HbA1c testing frequency in the decade following T2D diagnosis, and how this relates to all-cause mortality. By leveraging real-world primary care data from a large, representative UK patient population, this study aims to provide new insights into the role of mental health in diabetes management and its impact on all-cause mortality.

## Methods

### Data sources

This study used the Clinical Practice Research Datalink (CPRD) Aurum dataset, linked to the Hospital Episode Statistics Admitted Patient Care (HES APC) database. CPRD Aurum includes anonymised electronic health records (EHRs) from general practices across the UK using the EMIS software system, with records available for over 16 million current patients (17). It provides comprehensive records of clinical events, demographic measures, biochemical measures, and prescriptions. HES APC contains data from admissions to National Health Service (NHS) hospitals in England (18). Diagnoses in CPRD Aurum are recorded using READ or SNOMED-CT classifications. Diagnoses in HES APC are recorded using the International Classification of Diseases version 10 (ICD-10), with procedures coded using the UK Office of Population, Census and Surveys classification (OPCS).

### Study Population

Individuals with at least one T2D code between start of data collection (1988) and 31/12/2023 were identified from CPRD Aurum December 2023 data release (17) (Supplementary Table 1). A full description of the methods for T2D phenotype extraction is available in Figure 1 and online (19). Briefly, HbA1c values were extracted from CPRD Aurum using READ/SNOMED CT codes (Supplementary Table 2). Values which were below 20 were converted from DCCT units (%) to IFCC units (mmol/mol) and HbA1c values above 195 mmol/mol were removed as outliers. HbA1c values collected before 01/01/1990 were also excluded (20).

**Figure 1.**
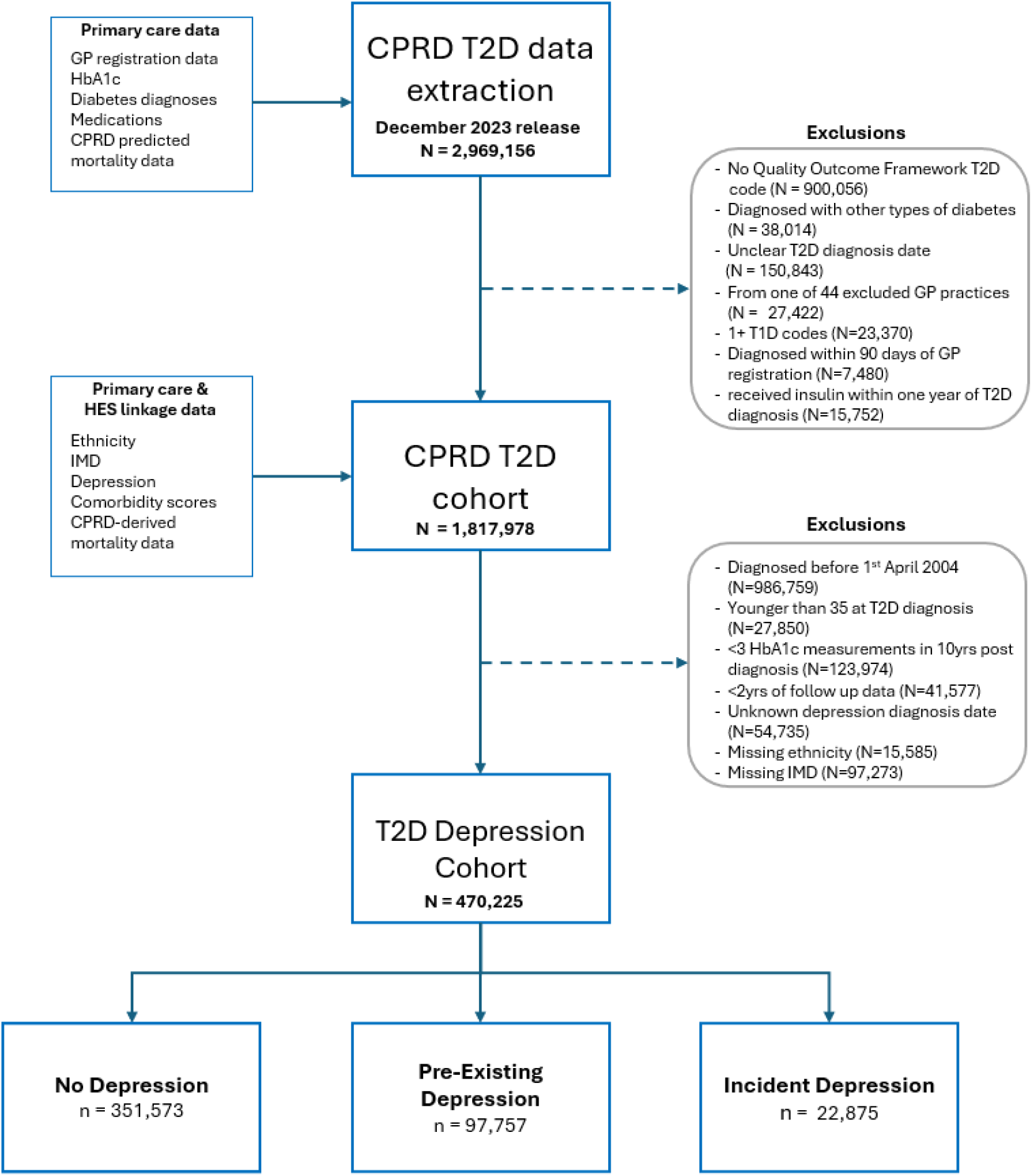
Overview of the exclusion and inclusion criteria used to define the depression cohorts. GP, general practice; CPRD, Clinical Practice Research Datalink; QCed, quality controlled; IMD, index of multiple deprivation; HbA1c, glycated haemoglobin; T1D, Type 1 diabetes; T2D, Type 2 diabetes.

Prescriptions for T2D medications were extracted using DM+D codes (Supplementary Tables 3 and 4). Individuals were included if they met the following criteria: 1) at least one Quality and Outcomes Framework (QOF) T2D code (Supplementary Table 5; n = 900,056 excluded), 2) no diagnosis of gestational diabetes, diabetes insipidus, Type 1 diabetes, or other rarer diabetes types (Supplementary Table 5; n = 53,889 excluded), 3) not registered at one of 44 excluded CPRD practices (n = 27,422 excluded), 4) diagnosed with T2D after the T2D QOF introduction (1^st^ April 2004; n = 986,759 excluded), and 5) diagnosed with T2D ≥90 days after registering with current practice (n = 7,480 excluded). T2D diagnosis date (index date) was calculated as the earliest of: (1) a broad T2D SNOMED code was recorded (excluding codes in year of birth), (2) an HbA1c ≥ 48mmol/mol (6.5%), or (3) prescription of an anti-diabetic medication. Individuals diagnosed with T2D before 35 years of age, or who were prescribed insulin within the first year after T2D diagnosis were excluded (n = 27,850 and n = 15,752, respectively).

### Outcomes

#### HbA1c testing frequency

HbA1c testing frequency was defined as the number of unique HbA1c test dates per year since T2D diagnosis. For years which were incomplete due to censoring or mortality, testing dates were counted to the censoring date. Individuals were followed from the T2D diagnosis date (index date) to the earliest of death, de-registration from CPRD Aurum practice, the practice’s last data collection date, 10 years from T2D diagnosis, or 31/12/2023. The maximum follow-up for this outcome was therefore 10 years. Individuals with fewer than three total HbA1c measurements, or who had HbA1c measurements in less than two years were excluded (n =123,974 and n = 41,577, respectively).

#### All-cause mortality

All-cause mortality was defined using CPRD’s algorithmically estimated date of death. Time-to-event was measured from the index date (T2D diagnosis) until death or censoring. Participants were censored at the earliest of the following: CPRD Aurum practice deregistration, last data collection date from the practice, or the study end date (31/12/2023). Follow-up was not restricted to 10 –years, unlike the HbA1c testing outcome.

### Exposures

Two depression phenotypes were considered as primary exposure variables, defined based on the relative timing of depression and T2D diagnoses. Individuals were defined as having depression if they had at least one depression READ3/SNOMED CT primary care code (Supplementary Table 7). The date of depression diagnosis was the date of the first recorded depression code. Those whose first depression code was historical, making the date of depression diagnosis ambiguous, were excluded (n = 54,735).

Two depression status exposure variables were then defined:

- **Pre-existing depression**. A binary variable indicating whether an individual had a recorded depression diagnosis at or before the date of their T2D diagnosis.
- **Incident depression**. A time-dependent binary variable that equals zero for all individuals until the date of their first recorded depression diagnosis after T2D diagnosis, after which it was set to one. Individuals with no recorded depression diagnosis during follow-up, including those with pre-existing depression, were zero for the entire study period.

In individuals with pre-existing depression, six additional exposures were considered to capture depression severity and recency: 1) total number of depression codes prior to T2D diagnosis, 2) time between first depression diagnosis and T2D diagnosis, 3) time since the last recorded depression code before T2D diagnosis and the index date, 4) recurrent pre-existing depression, defined as having more than one date with a depression code prior to T2D diagnosis, 5) first diagnosis of depression within 10 years of T2D diagnosis, and 6) last depression code within 10 years before T2D diagnosis.

### Covariates

#### Demographic characteristics

Gender was defined according to self-reported gender available in the CPRD. Age at T2D diagnosis was calculated as the difference between date of birth and the index date. Ethnicity was defined using self-reported responses to the 16-category NHS ethnicity classification. A binary variable for ethnicity was created, identifying individuals who self-reported any code containing “White”, and those that did not. Due to the limited sample size within specific ethnicity strata, we did not stratify these analyses by ethnicity. Patient-level Index of multiple deprivation (IMD) deciles and year of T2D diagnosis were also included as covariates for all analyses. Due to limited computational resources, IMD was treated as a continuous variable rather than a categorical variable, where a higher IMD score represents higher deprivation. As this study used complete case analysis, individuals with missing covariate data were removed (n = 15,585 and 92,723 for ethnicity and IMD, respectively).

#### Comorbidity scores

The following 13 comorbidities were extracted from CPRD, HES APC and OPCS-4: frailty (using direct primary care coding), heart failure, chronic liver disease, dementia, diabetic nephropathy, ischemic heart disease (excluding myocardial infarction), myocardial infarction, diabetic neuropathy, peripheral artery disease, diabetic retinopathy, trans-ischaemic aneurysm, ischaemic and haemorrhagic stroke (excluding trans-ischaemic aneurysm), and stage V chronic kidney disease (CKD; also known as end-stage renal disease) (Supplementary Tables 8, 9, and 10). CKD status was also determined based on estimated glomerular filtration rate (eGFR). Briefly, eGFR codes were extracted from the primary care, and creatinine-based eGFR was estimated using the creatinine EPI 2021 equation (Supplementary Table 11). Individuals were considered to have CKD if they had at least two consecutive eGFR at least 90 days apart which were below 60ml/min/1.73m^2^ and would therefore meet the diagnostic criteria for stage III CKD (21). A comorbidity was considered as pre-existing if it was diagnosed before T2D, and incident if the comorbidity was first diagnosed after T2D diagnosis. Two comorbidity scores were then defined. A pre-existing comorbidity score was constructed as the sum of the number of comorbidities diagnosed at or before T2D diagnosis, and an incident comorbidity score was calculated as the time-varying cumulative sum of comorbidities diagnosed after T2D diagnosis.

#### Healthcare use in primary care

Healthcare use in primary care was defined as the number of unique patient-date combinations on which any primary care code (excluding prescription codes) was recorded in CPRD (22). To avoid double counting, dates on which HbA1c testing occurred were excluded.

### Statistical analysis

#### Baseline characteristics

Baseline characteristics were summarised according to depression status (no depression vs pre-existing depression vs incident depression) and compared across the depression groupings using likelihood ratio tests for dichotomous variables, and ANOVA for continuous variables.

#### The association between exposures and HbA1c testing frequency

We used Conway-Maxwell-Poisson mixed effects models (CMP-MEMs) to investigate the association between the depression exposures and the annual HbA1c testing frequency in primary care over time since T2D diagnosis. For follow-up years truncated by death or censoring, the log of the fraction of the year under observation was included as an offset. CMP-MEMs were selected to accommodate the under-dispersion observed in HbA1c testing frequency. All models included a natural spline with three degrees of freedom to allow for the non-linear time effects. A random intercept was included to capture between-subject variation. A model with a random slope for time was tested, but its variance was near zero, suggesting that the added complexity was unnecessary.

Three sequential models were fitted, each adding additional fixed effects covariates. Model A included only demographic covariates: age at diagnosis, gender, a binary ethnicity variable, year of T2D diagnosis, and IMD decile. Model B added the depression exposure of interest. Model C further included pre-existing and incident comorbidity scores. Effect sizes from CMP-MEMs are reported as incident rate ratios (IRRs) with 95% confidence intervals, representing how HbA1c testing frequency changes per unit increase in a continuous exposure variable (e.g. comorbidity score), or per category change in dichotomous variables (e.g. pre-existing or incident depression).

#### The association between exposures and all-cause mortality

We applied Cox proportional hazards models to estimate the association of the exposure variables with all-cause mortality, reporting hazard ratios (HRs) with 95% confidence intervals (CIs). HbA1c testing frequency was included as a time-varying covariate by counting the number of measurement dates recorded within yearly time intervals, from the year prior (lagged values). Initial models assessed HbA1c testing frequency as a continuous variable; however, evidence of non-linearity led us to use four categories (0, 1– 2, 3–4, ≥5 tests per year), with no tests per year as the reference group. To mitigate potential bias due to excluding individuals with insufficient HbA1c measurements, we applied a landmark design, with individuals entering the risk set at two years post-T2D diagnosis, provided they were alive and under observation. This ensured all individuals had sufficient opportunity to accumulate HbA1c measurements prior to entering the risk set for all-cause mortality.

To determine the effect of the HbA1c testing frequency categories on all-cause mortality, three models were fitted. The first model included only demographic covariates and the depression exposure of interest. In the second model, the HbA1c testing categories were added, and the final model also included comorbidity scores. We assessed proportional hazards assumptions for key covariates (pre-existing depression, incident depression, baseline comorbidity score, incident comorbidity score, HbA1c testing frequency categorical variable using the Grambsch-Therneau test (23). Where violations were detected, and to explore how covariate effects changes over time, we fitted time-stratified Cox models across four follow-up intervals: two to five years, five to eight years, and eight to eleven years, and eleven to fourteen years following T2D diagnosis. Given the large sample size and model complexity, incorporating time interaction terms to accommodate the non-proportional hazards assumption was not feasible. A Bonferroni-corrected multiple testing threshold of 6.3×10^−3^ (0.05/8) was applied to account for the largest number of tests in a single analysis section.

Since each primary care observation provides an opportunity for HbA1c measurement, we wished to determine whether the results of our study were driven by increased engagement with primary care overall, rather than a specific effect of increased HbA1c testing. As a sensitivity analysis, we examined the association of primary care healthcare use with all-cause mortality. Given the strong correlation between the incident comorbidity score and healthcare use, the pre-existing and incident comorbidity scores were omitted in these models. We compared the association of HbA1c testing frequency categories with all-cause mortality when adjusting for healthcare use instead of comorbidity, alongside demographic covariates.

#### Materials

All analyses were performed using R v.4.3.2. Data extraction and preprocessing were performed using the Exeter Diabetes CPRD analysis package (24). CMP-MEMs were fitted using the *glmmTMB* package (25). Cox proportional hazards models and related sensitivity analyses were performed with the s*urvival* package (23). Figures were generated using the *ggplot2* package (26).

#### Code and data availability

All anonymised code is available at https://github.com/Exeter-Diabetes/CPRD-Dale-MDD-HbA1c-testing. This project was conducted under CPRD study reference ID 23_002544. Researchers wishing to access CPRD data must obtain the necessary approvals and data access agreements.

## Results

### Cohort description

A total of 470,225 individuals met all study criteria and were included in the analysis (Table 1). The median follow-up time for mortality was 8.6 years, and 35.2% of individuals included had 10 years of HbA1C measurements available. Males comprised 56.2% (n = 246,066) of the cohort and 82.4% self-reported their ethnicity as “White” (n = 398,440). The median age at T2D diagnosis (index date) was 61 years. At diagnosis, 36% of individuals had at least one comorbidity (n = 168,421), which increased to 80.9% by the end of the mortality analysis follow-up (n = 378,002; Supplementary Figures 1, 2, and 3).

**Table 1.**
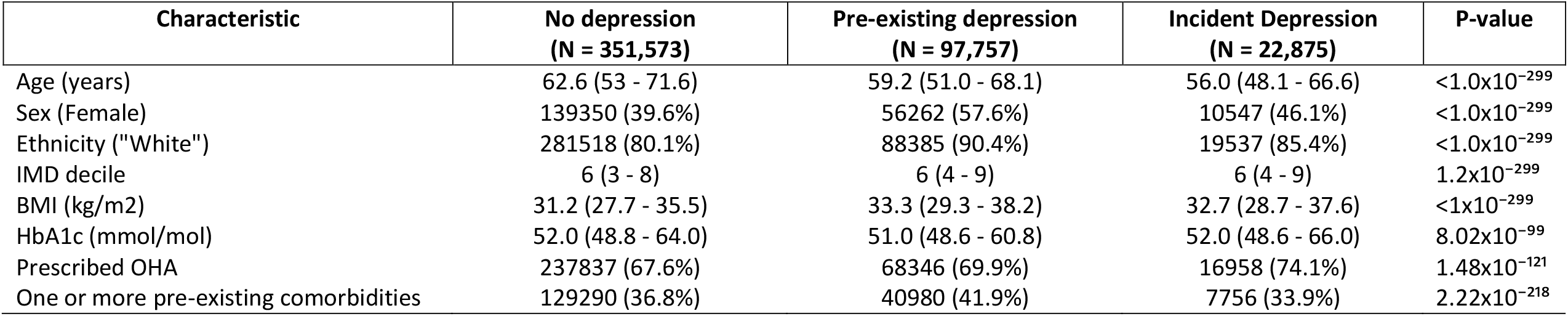
Clinical and demographic characteristics at type 2 diabetes diagnosis, stratified by depression status. Median and IQR are reported for continuous variables, and number of individuals and percentage for dichotomous variables. IMD, index of multiple deprivation; BMI, body mass index; OHA, Oral hypoglycaemic agents. “Prescribed OHA” refers to receiving an OHA prescription within one year of T2D diagnosis. p-values represent testing differences in characteristics between all groups.

| Characteristic | No depression<br>(N = 351,573) | Pre-existing depression<br>(N = 97,757) | Incident Depression<br>(N = 22,875) | P-value |
| --- | --- | --- | --- | --- |
| Age (years) | 62.6 (53 - 71.6) | 59.2 (51.0 - 68.1) | 56.0 (48.1 - 66.6) | <1.0x10<sup>-299</sup> |
| Sex (Female) | 139350 (39.6%) | 56262 (57.6%) | 10547 (46.1%) | <1.0x10<sup>-299</sup> |
| Ethnicity ("White") | 281518 (80.1%) | 88385 (90.4%) | 19537 (85.4%) | <1.0x10<sup>-299</sup> |
| IMD decile | 6 (3 - 8) | 6 (4 - 9) | 6 (4 - 9) | 1.2x10<sup>-299</sup> |
| BMI (kg/m2) | 31.2 (27.7 - 35.5) | 33.3 (29.3 - 38.2) | 32.7 (28.7 - 37.6) | <1x10<sup>-299</sup> |
| HbA1c (mmol/mol) | 52.0 (48.8 - 64.0) | 51.0 (48.6 - 60.8) | 52.0 (48.6 - 66.0) | 8.02x10<sup>-99</sup> |
| Prescribed OHA | 237837 (67.6%) | 68346 (69.9%) | 16958 (74.1%) | 1.48x10<sup>-121</sup> |
| One or more pre-existing comorbidities | 129290 (36.8%) | 40980 (41.9%) | 7756 (33.9%) | 2.22x10<sup>-218</sup> |

Pre-existing depression was present in 20.1% (*N* = 97,637) of the cohort. In this subgroup, the median interval between last recorded depression code and T2D diagnosis was nine years, and 63.6% of these individuals had recurrent depression prior to T2D diagnosis. Of the individuals without a pre-existing depression diagnosis, 22,875 individuals (6.1%) developed depression following T2D, with a median time to depression of 5.32 years. Compared to individuals with no depression, individuals with any depression were younger, more likely to be female, more likely to be “White”, and had higher deprivation. They also had higher BMI at T2D diagnosis and were more likely to be prescribed an oral hypoglycaemic agent within one year of T2D diagnosis (p < 6.3×10^−3^ for all reported results) (Table 1).

### Depression status is associated with increased annual HbA1c testing frequency

HbA1c testing was most frequent during the first year after T2D diagnosis, with a median of 2.35 HbA1c measurements. This frequency decreased to 1.45 measurements in the second year and then stabilized, reaching 1.41 by the tenth-year post-diagnosis. The median time between tests was 186 days. Individuals with pre-existing or with incident depression followed a similar early pattern but had more frequent testing than those without depression after the second year and remained higher until 10 years after diagnosis. (Supplementary Figures 5, and 6).

Results from Model A, including only demographic covariates, showed that older age at T2D diagnosis (IRR: 1.05, CI: 1.04 – 1.06, p = <1×10^−200^), and self-reported White ethnicity were associated with higher HbA1c testing frequency in the decade following T2D diagnosis, whereas being female was associated with lower testing (IRR: 0.92, CI: 0.91 – 0.93, p = <1×10^−200^) (Table 2). Higher deprivation (IMD decile) was also linked to a modest increase in testing (IRR: 1.00, 95% CI: 1.00 – 1.01, p = 5.4×10^−111^).

**Table 2.**
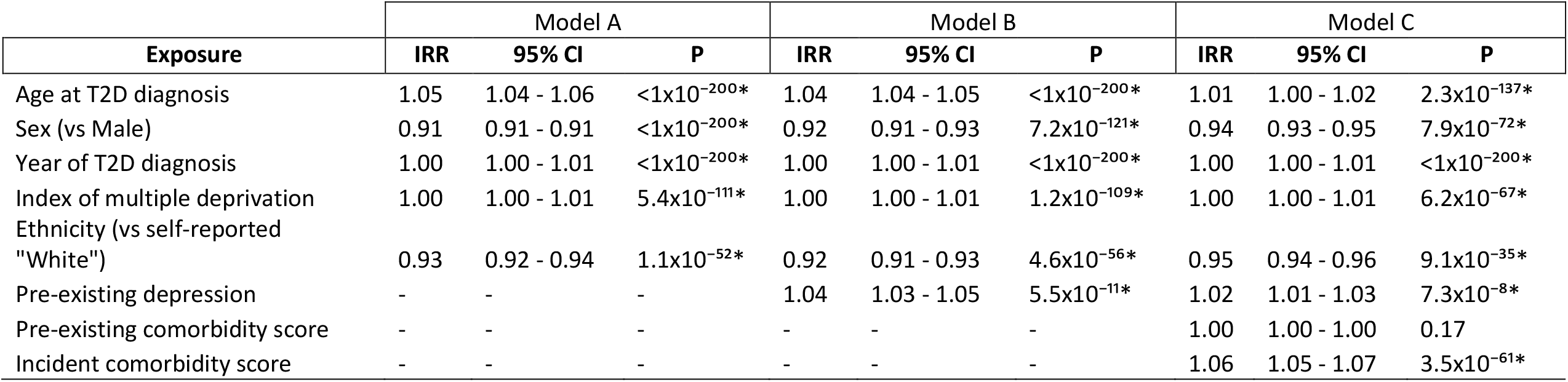
Association of pre-existing depression with HbA1c testing frequency, from fixed effect analysis. Effects sizes and error are presented as incident rate ratio (IRR) and 95% confidence intervals (CI). Model A, model including covariates; Model B, model including covariates and depression status; Model C, model including covariates, depression status, and comorbidity scores; *, statistically significant after Bonferroni multiple testing correction (p < 6.3×10^−3^).

**Table 3.** the association of incident depression with HbA1c testing frequency from the fixed effect analysis. Effects sizes and error are presented as incident rate ratio (IRR) and 95% confidence intervals, respectively. IRR, incidence rate ratio; CI, 95% confidence intervals. Model A, model including covariates; Model B, model including covariates and depression status; Model C, model including covariates, depression status, and comorbidity scores; *, statistically significant after Bonferroni multiple testing correction (p < 6.3×10^−3^).

| Exposure | Model A |  |  | Model B |  |  | Model C |  |  |
| --- | --- | --- | --- | --- | --- | --- | --- | --- | --- |
|  | IRR | 95% CI | P | IRR | 95% CI | P | IRR | 95% CI | P |
| Age at T2D diagnosis | 1.05 | 1.04 - 1.06 | $<1 \times 10^{-200*}$ | 1.04 | 1.04 - 1.05 | $<1 \times 10^{-200*}$ | 1.01 | 1.00 - 1.02 | $5.3 \times 10^{-144*}$ |
| Sex (vs Male) | 0.92 | 0.92 - 0.92 | $<1 \times 10^{-200*}$ | 0.92 | 0.91 - 0.93 | $1.1 \times 10^{-127*}$ | 0.93 | 0.92 - 0.94 | $7.9 \times 10^{-81*}$ |
| Year of T2D diagnosis | 1.00 | 1.00 - 1.01 | $<1 \times 10^{-200*}$ | 1.00 | 1.00 - 1.01 | $<1 \times 10^{-200*}$ | 1.00 | 1.00 - 1.01 | $<1 \times 10^{-200*}$ |
| Index of multiple deprivation | 1.00 | 1.00 - 1.01 | $7.3 \times 10^{-118*}$ | 1.00 | 1.00 - 1.01 | $1.2 \times 10^{-106*}$ | 1.00 | 1.00 - 1.01 | $8.4 \times 10^{-73*}$ |
| Ethnicity (vs self-reported "White") | 0.93 | 0.92 - 0.94 | $5.9 \times 10^{-58*}$ | 0.92 | 0.91 - 0.95 | $7.6 \times 10^{-54*}$ | 0.95 | 0.94 - 0.96 | $9.1 \times 10^{-33*}$ |
| Incident depression | - | - | - | 1.06 | 1.05 - 1.07 | $5.5 \times 10^{-18*}$ | 1.04 | 1.03 - 1.05 | $5.5 \times 10^{-11*}$ |
| Pre-existing comorbidity score | - | - | - | - | - | - | 1 | 1.00 - 1.00 | 0.11 |
| Incident comorbidity score | - | - | - | - | - | - | 1.07 | 1.06 - 1.08 | $6.1 \times 10^{-66*}$ |

Adding depression exposures (Model B) showed both incident (IRR: 1.06, 95% CI: 1.05 – 1.07, p = 6.5×10^−18^) and pre-existing depression (IRR: 1.04, 95% CI: 1.03 – 1.05, p = 4.7×10^−11^) were associated with more frequent HbA1c testing.

When the comorbidity scores were included (Model C), a higher incident, but not pre-existing, comorbidity score was associated with increased HbA1c testing. The associations of both depression exposures were attenuated but remained statistically significant (incident depression IRR: 1.04, 95% CI: 1.03 – 1.05, p = 5.5×10^−11;^ pre-existing depression IRR: 1.02, 95% CI: 1.01 – 1.03, p = 7.3×10^−8^).

### Increased HbA1c testing frequency is associated with decreased risk of all-cause mortality

In the full analysis cohort (N = 470,225; 4,294,629 person-years), the mortality rate was 196 per 10,000 person-years, with death occurring in 17.8% of participants during follow-up (for depression stratified mortality rates, see Supplementary Table 12).

Older age at T2D diagnosis (HR: 1.07, 95% CI: 1.07 – 1.07, p = <1×10^−256^) and higher deprivation (HR: 1.04, 95% CI: 1.04 – 1.04, p = 1.1×10^−212^) were associated with higher mortality, whereas female sex (HR: 0.83, 95% CI: 0.81 – 0.84, p = 2.1×10^−152^) and self-reported non-White ethnicity (HR: 0.70, 95% CI: 0.68–0.72, p = 8.2×10^−153^) were associated with reduced mortality risk. Higher comorbidity scores were also associated with an increased risk of all-cause mortality, with the effect being greater for the incident (HR: 1.37, 95% CI: 1.37 – 1.38, p = <1×10^−256^) compared with the pre-existing comorbidity score (HR: 1.30, 95% CI: 1.29 – 1.30, p = <1×10^−256^).

Both one to two HbA1c tests/year and three to four tests/year were associated with lower mortality when compared with no tests (HR: 0.80, 95% CI: 0.78 – 0.81, p = 1.9×10^−107^; HR: 0.77, 95% CI: 0.75 – 0.79, p = 2.38×10^−84^). However, the risk of all-cause mortality in patients tested 5+ time per year was not statistically different from that in patients with no HbA1c testing (HR: 0.92, 95% CI: 0.85 – 1.00, p = 0.05) (Figure 2).

**Figure 2.**
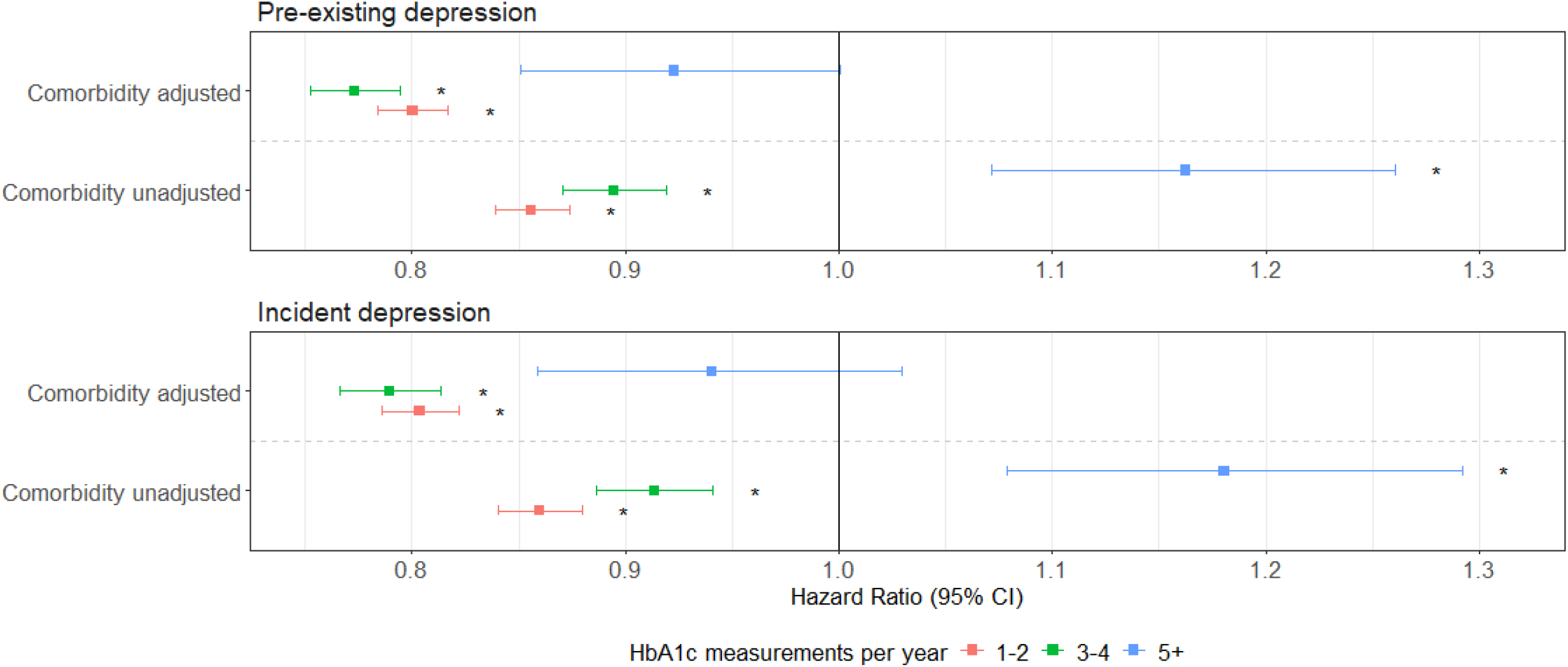
Forest plots for the associations between HbA1c testing frequency categories and risk of all-cause mortality. Comorbidity unadjusted, model including covariates and depression status; Comorbidity adjusted, model including covariates, depression status, and pre-existing and incident comorbidity scores. Error bars represent 95% confidence intervals. *, statistically significant after Bonferroni multiple testing correction (p < 6.3×10^−3^).

### Depression status is associated with increased risk of all-cause mortality

In models only adjusting for demographic characteristics, both pre-existing and incident depression status were associated with higher mortality risk (pre-existing depression HR: 1.19, 95% CI: 1.17 – 1.21, p = 6.6×10^−82^; incident depression HR: 1.41, CI: 1.37 – 1.46, p = 3.7×10^−97^). Adjusting for categorical HbA1c testing frequency did not meaningfully change the size of these associations (Figure 3). After adjusting for the comorbidity scores, effect sizes were substantially attenuated but remained statistically significant (pre-existing depression HR: 1.05, 95% CI: 1.03 – 1.07, p: 9.6×10^−8^; incident depression HR: 1.19, 95% CI: 1.15 – 1.22, p = 1.1×10^−24^) (Figure 3).

**Figure 3.**
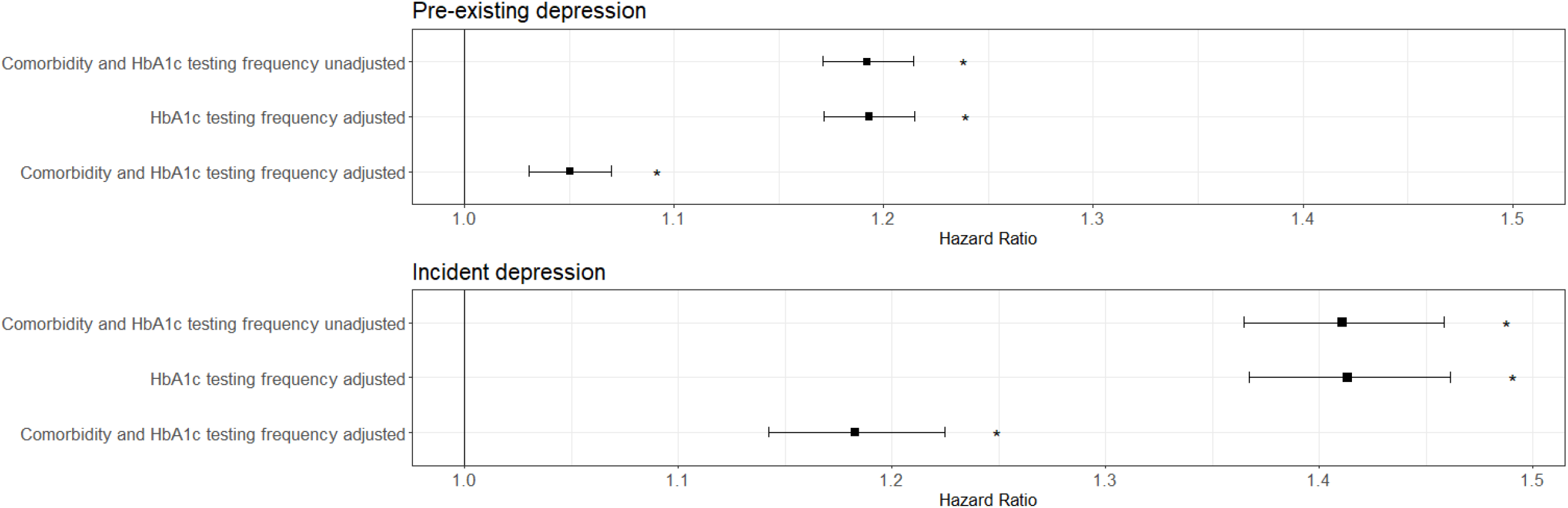
Forest plots for the associations between depression status and risk of all-cause mortality. Error bars represent 95% confidence intervals. All models include covariates and depression status. Comorbidity adjusted, model includes pre-existing and incident comorbidity scores; *, statistically significant after Bonferroni multiple testing correction (p < 6.3×10^−3^).

However, several key covariates (HbA1c testing frequency, the comorbidity scores and pre-existing depression status) violated the proportional hazards assumption (Supplementary Table 13). Time-stratified Cox models demonstrated that associations varied across follow-up intervals. For HbA1c testing frequency, taking the effect of having one to two HbA1c tests/year compared to no tests as an example, the protective association was weaker early in follow-up (HR = 0.92, 95% CI: 0.88 – 0.96 in years two to five) but became stronger in later periods (HR = 0.70, 95% CI: 0.66 - 0.74 in years five to eight, remaining similar thereafter) (Supplementary Figure 7).

Both pre-existing and incident comorbidity scores were consistently associated with higher mortality, but effect sizes attenuated slighted with longer follow-up (Supplementary Figures 8 and 9). For the pre-existing comorbidity score, HRs declined from 1.31 (95% CI: 1.28 – 1.33) in years two to five to 1.26 (95% CI: 1.23 – 1.28) in years 11-14. For the incident comorbidity score, the HR was 1.44 (95% CI: 1.42 – 1.46) in years two to five, reducing to 1.32 (95% CI: 1.31 – 1.33) by years 11–14. For pre-existing depression, the HR was 1.07 (95% CI: 1.03 – 1.11) in years two to five. This declined to 1.00 (95% CI: 0.96 - 1.04) in years 8-11, before increasing again to 1.07 (95% CI: 1.02 – 1.13) between 11 −14 years (Supplementary figure 10). For incident depression, the effect on mortality was strongest during the first two to five years following T2D diagnosis (HR: 1.28, 95% CI: 1.19 – 1.38) but was reduced during years five to eight (HR: 1.14, 95% CI: 1.06 - 1.22) and remained similar in subsequent follow-up periods (Supplementary Figure 11).

### Pre-existing depression subgroup analyses

For both outcomes, annual HbA1c testing frequency over time since T2D diagnosis and all-cause mortality, we performed a subgroup analysis focusing on participants with pre-existing depression (N = 97,757). In the fully adjusted models including comorbidity scores, none of the depression severity variables were associated with a statistically significant change in HbA1c testing frequency after multiple testing correction (Supplementary figure 12). While recurrent depression status (HR: 1.05, 95% CI = 1.01 – 1.09, p = 3.2×10^−4^) and having a depression clinical code within 10 years of diagnosis (HR: 1.06, 95% CI = 1.04−1.12, p = 7.8×10^−6^) were associated with high risk of all-cause mortality, and having a longer duration between depression diagnosis and T2D diagnosis was associated with a lower risk of all-cause mortality (HR: 0.99, 95%CI = 0.99 – 1.00, p = 1.1×10^−3^), none of these associations remained statistically significant after adjusting for comorbidity scores (Supplementary Figures 13 and 14).

### Sensitivity Analysis: healthcare use

Given the plausible link between healthcare use and HbA1c testing frequency, we tested whether primary care healthcare use was driving the association between increased HbA1c testing frequency and all-cause mortality (supplementary figures 15 and 16). When included in the demographic covariate model, higher healthcare use was strongly associated with increased risk of all-cause mortality (HR = 1.02, 95% CI = 1.02 − 1.02, p = < 1×10^−299^). Compared to the model adjusted for covariates and depression, adjusting for healthcare use increased the effect size of one to two HbA1c tests/year (HR: 0.77, 95% CI: 0.75 – 0.79, p = 1.4×10^−116^), and three to four tests/year (HR: 0.70, 95% CI = 0.68 – 0.72, p = 2.9×10^−124^) for all-cause mortality. The direction of effect for five or more tests/year was reversed healthcare use unadjusted model and having five or more tests/year was associated with lower all-cause mortality (HR: 0.76, 95% CI = 0.69 − 0.83, p = 1.5×10^−9^) (Supplementary figure 17).

## Discussion

This study used primary care EHRs from the CPRD to examine the relationship between pre-existing and incident depression, HbA1c testing frequency and all-cause mortality in individuals with T2D. We found that both pre-existing, and incident depression were associated with increased HbA1c testing frequency in the decade following T2D diagnosis, yet this did not translate into a lower risk of mortality. Instead, both depression exposures were associated with an increased risk of all-cause mortality, even after accounting for comorbidities, whilst higher glycaemic testing displayed a non-linear association with a reduced mortality risk.

The positive association between depression and glycaemic testing is somewhat unexpected given that depression is typically associated with lower diabetes-related self-testing and reduced medication adherence (14,15), but reflect both clinical practice and the interplay between depression and diabetes severity Clinicians may increase monitoring for individuals with depression due to concerns about self-care and overall health. Additionally, poorer self-perceived health and the accumulation of comorbidities, which may reflect more severe T2D, are linked to an increased risk of developing depression (27,28). Conversely, having a depression diagnosis is associated with poorer glycaemic control and adverse health behaviours, such as reduced physical activity, poor diet and lower engagement with self-care behaviours, all of which contribute to faster T2D progression (15,29–31). NICE guidelines recommend more frequent testing for individuals with more severe diabetes, poorer overall health and declining glycaemic control (2). This bidirectional relationship, whereby poor health increases depression risk and depression worsen diabetes outcomes, may therefore lead to higher testing frequency. Consistent with this, adjusting for comorbidity scores attenuated the association between depression (both pre-existing and incident) and glycaemic testing.

In addition to planned HbA1c measurements, studies of overall healthcare utilisation have found that individuals with depression tend to have more frequent contact with healthcare services, which may lead to increased opportunistic testing (16). However, this does not explain why individuals with depression attend these appointments, despite depression’s association with reduced engagement in diabetes self-care (15). In this study, higher healthcare use was associated with increased risk of all-cause mortality, and adjusting for primary care healthcare use strengthened the protective association between HbA1c testing frequency and all-cause mortality. This finding suggests that more frequent testing, while correlated with overall healthcare use, likely captures a different aspect of T2D management than overall healthcare use alone which should be investigated further.

The mortality benefit of HbA1c monitoring was confined to moderate testing frequencies (1–4 tests per year). Individuals with 5 or more tests annually did not experience the same survival advantage, and after adjustment for comorbidity burden the association was null. This likely reflects the fact that very frequent testing is concentrated among patients with greater illness severity or unstable glycaemic control, where closer monitoring is a response to clinical complexity rather than an independent protective factor. These findings reinforce the view that testing frequency serves as a marker of disease severity and healthcare needs, rather than a direct mechanism influencing survival. Consequently, we propose that longitudinal HbA1c testing patterns may serve as a useful indicator for identifying clinically vulnerable patients.

Another key consideration is how depression was identified in this study. Individuals in our cohort were required to have received a primary care code for depression, meaning that those who sought medical attention for their mental health were captured. This introduces potential surveillance bias and limits generalisability to undiagnosed cases, who may have different healthcare-seeking behaviours. Many studies have demonstrated the effect of depression on other metrics of diabetes self-care and increased risk of mortality. However, no studies have looked at HbA1c testing frequency and investigated the impact of depression on mortality in T2D while accounting for testing patterns. Frequent glycaemic testing is important for maintaining optimal glycaemic control, which in turn reduces the risk of complications and all-cause mortality (2). Therefore, our finding that increased HbA1c measurement frequency was associated with a lower risk of all-cause mortality aligns with expectations. However, despite depression being associated with increased testing, it remains associated with a higher risk of mortality. This may be through mechanisms such as lower medication adherence (32) and broader health behavioural pathways (diet, smoking, exercise, etc) as well as chronic inflammation (9) and HPA axis dysregulation (12). These may not be fully mitigated through testing, as testing provides an opportunity for increased glycaemic control as a partnership between the patient and physician, not a guarantee of improvement in glycaemic control.

This study has several strengths. First, it benefits from a large and representative sample of individuals with T2D receiving primary care in England, supporting generalisability of our findings to this population. our study includes follow up for up to 18 years after T2D diagnosis for the all-cause mortality analyses, allowing us to assess the long-term impact of depression on HbA1c testing and mortality. Second, in contrast to many studies that treat depression as a single dichotomous variable, our stratification by timing and severity enabled us to assess how both the time of onset and severity of depression influence HbA1c testing frequency and all-cause mortality risk. The use of longitudinal modelling allowed us to characterise temporal relationships between depression, HbA1c testing frequency, and mortality more accurately than would be possible in a cross-sectional design. Additionally, our careful evaluation of the proportional hazards assumption in the Cox models provides confidence in the robustness of the observed associations.

Several limitations should also be acknowledged. First, findings may not generalize beyond the English primary care setting. Second, some HbA1c measurements included in the analysis were taken less than two months apart. These are more likely to be opportunistic measurements and may overestimate testing frequency relevant for diabetes care. In fact, while our analysis suggests at least 9% of all tests may have arisen from opportunistic testing (given they were within two months of a previous test), adjustment for either comorbidity scores or primary care healthcare use greatly increased the effect size of HbA1c testing on all-cause mortality, which suggests that the pleiotropic effect of opportunistic testing on all-cause mortality may be captured by adjusting for proxies of overall health such as comorbidity status and healthcare use in our analysis. Third, changes in diagnostic criteria for T2D over time and the assumption of linear covariates effects (for example, with the comorbidity scores) could contribute to residual confounding. In addition, the restructuring of time into annual intervals for counting HbA1c measurements may introduce some measurement imprecision. Finally, comorbidity scores may act as both mediators and confounders in the depression-diabetes relationship, complicating interpretation.

## Conclusion

Individuals with depression receive more frequent HbA1c testing but still experience higher all-cause mortality. While higher testing was independently associated with reduced mortality risk, it did not mitigate the elevated risk observed for those with depression. These findings suggest that how individuals with depression interact with primary care for T2D glycaemic monitoring is not a primary driver of the increased mortality risk observed in individuals with depression and T2D. Integrating depression management and supporting medication adherence, alongside HbA1c testing, are likely necessary to reduce mortality in people with comorbid T2D and depression. Future research should investigate additional metrics of glycaemic testing, such as adherence to HbA1c testing guidelines, as well as measures of depression severity, to discern the full relationship between HbA1c testing and mortality in individuals with co-morbid T2D and depression.

## Acknowledgements and funding

This work was funded by MRC project grant MR/X009815/1. This study was supported by the National Institute for Health and Care Research Exeter Biomedical Research Centre and by the NIHR Maudsley Biomedical Research Centre at South London and Maudsley NHS Foundation Trust and King’s College London. The views expressed are those of the author(s) and not necessarily those of the NIHR or the Department of Health and Social Care.

## Manuscript contributions

DH contributed to the conception of the study, performed the analysis, and prepared and revised the manuscript. KGY contributed to data preparation and management and revised the manuscript. RB and JT contributed to the study’s conception and revised the manuscript. CWHL provided guidance on the study’s direction and revised the manuscript. CML contributed to the study conception, assisted in manuscript preparation and revision, and provided guidance on the study’s direction. ACG contributed to the conception of the study, provided guidance for the study’s direction, and prepared and revised the manuscript.

## Guarantor statement

D.H. is the guarantor of this work and, as such, had full access to all the data in the study and takes responsibility for the integrity of the data and the accuracy of the data analysis.

## Ethics statement

This research has been conducted using the Clinical Practice Research Datalink under protocol 23_002544. Ethical approval for observational studies using the Clinical Practice Research Datalink is granted by the NHS Health Research Authority (Derby Research Ethics Committee; REC reference 21/EM/0265). Individual patient consent is not required.

