## Supplementary Figures for "Depression increases both glycated haemoglobin testing frequency and risk of all-cause mortality in type 2 diabetes"

Supplementary figure 1. Distribution of pre-existing comorbidities, stratified by depression status. Error bars represent 95% confidence intervals.

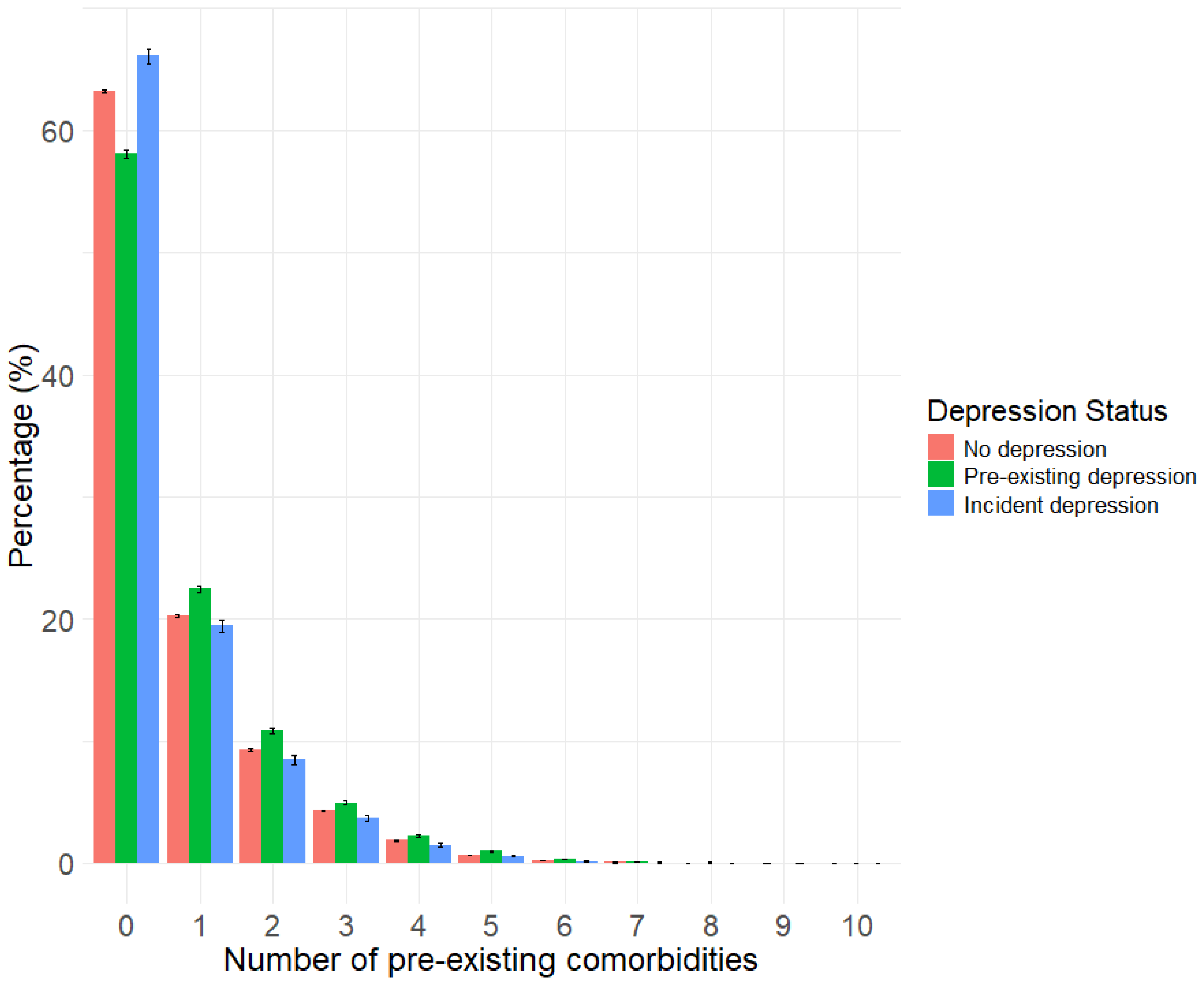

Supplementary Table 2. Number of incident comorbidities, stratified by depression status.

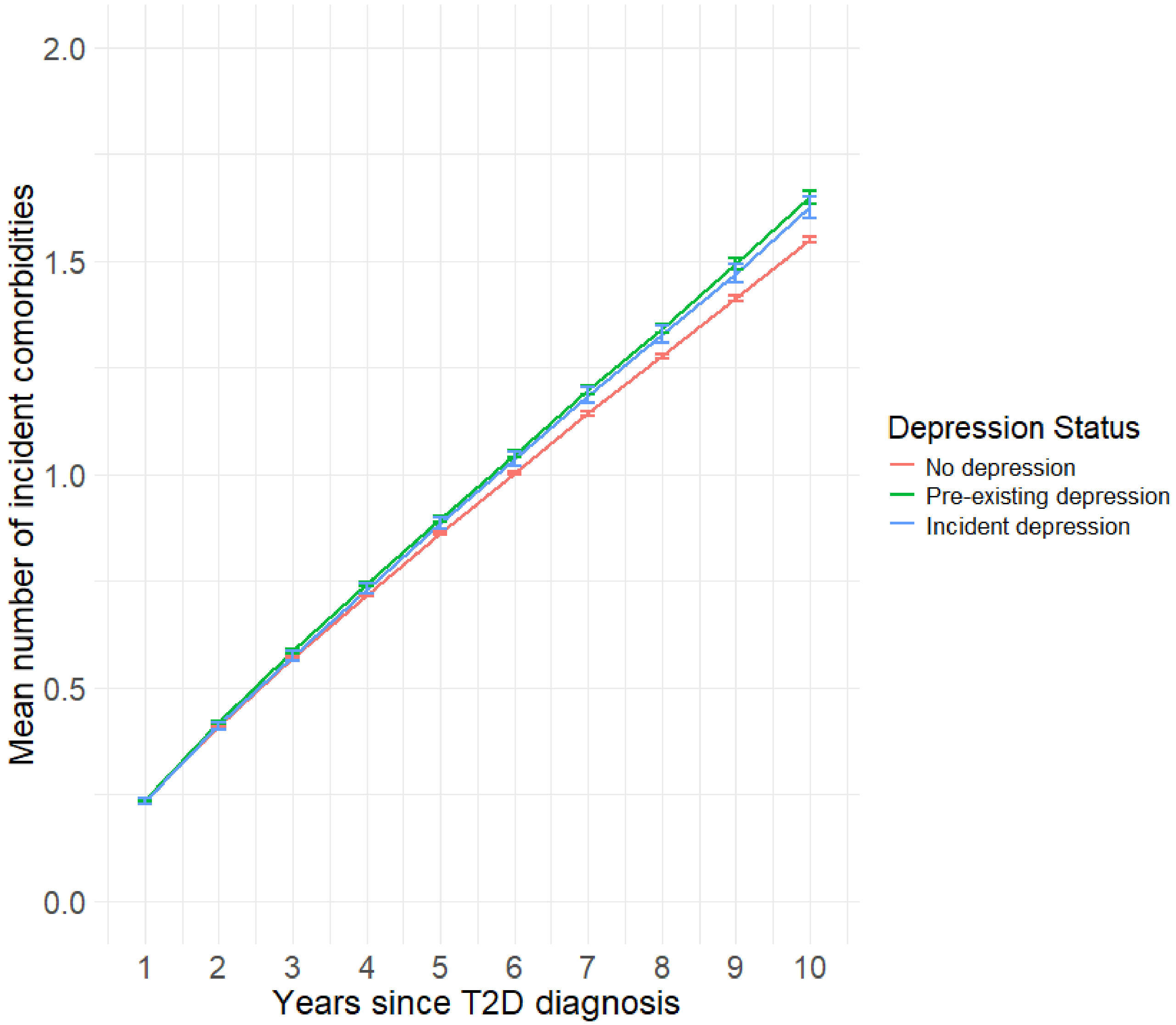

Supplementary figure 3. Accumulation of incident comorbidities across the first ten years of T2D diagnosis, stratified by time between T2D diagnosis and depression diagnosis, compared to individuals who do not have a pre-existing depression diagnosis and never receive a diagnosis of incident depression. Error bars represent 95% confidence intervals. Dashed lines represent the time window in which individuals in the incident depression group received a diagnosis.

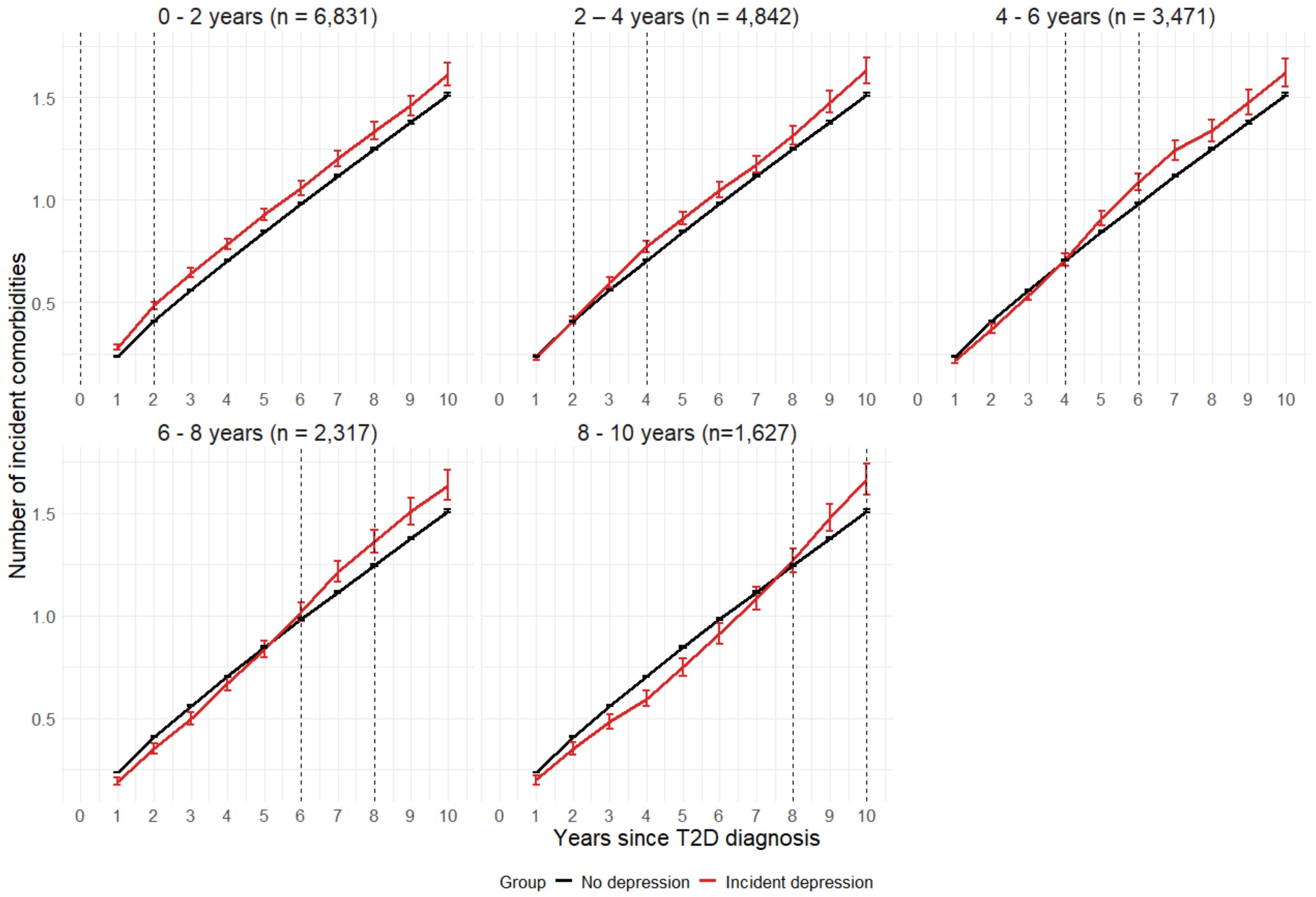

Supplementary Figure 4. Histogram of days between consecutive HbA1c tests. Error bars represent 95% confidence intervals. Vertical lines represent 60 days and 90 days cutoffs for potentially uninformative HbA1c testing.

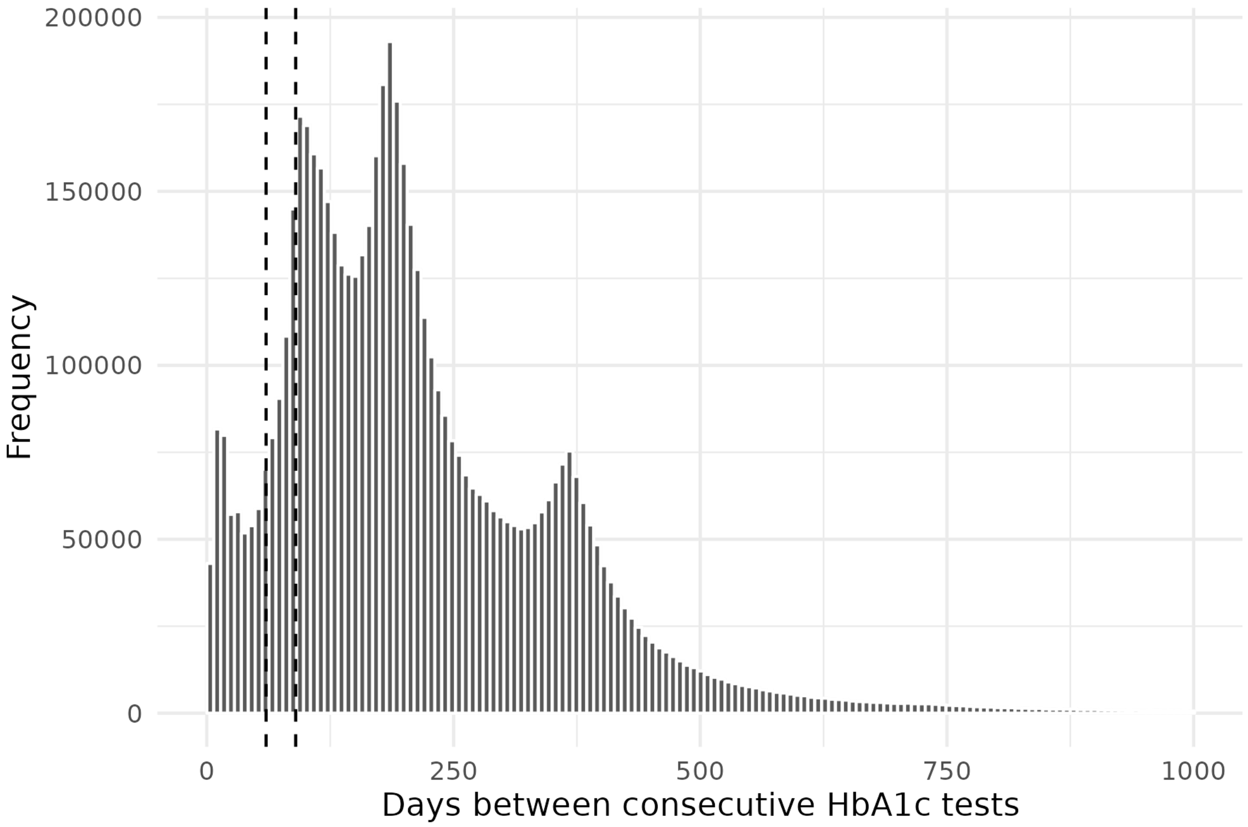

Supplementary Figure 5. mean HbA1c monitoring frequency by year from T2D diagnosis, stratified by depression status. Incident depression represents ever receiving a diagnosis of incident depression during follow up. Error bars represent 95% confidence intervals.

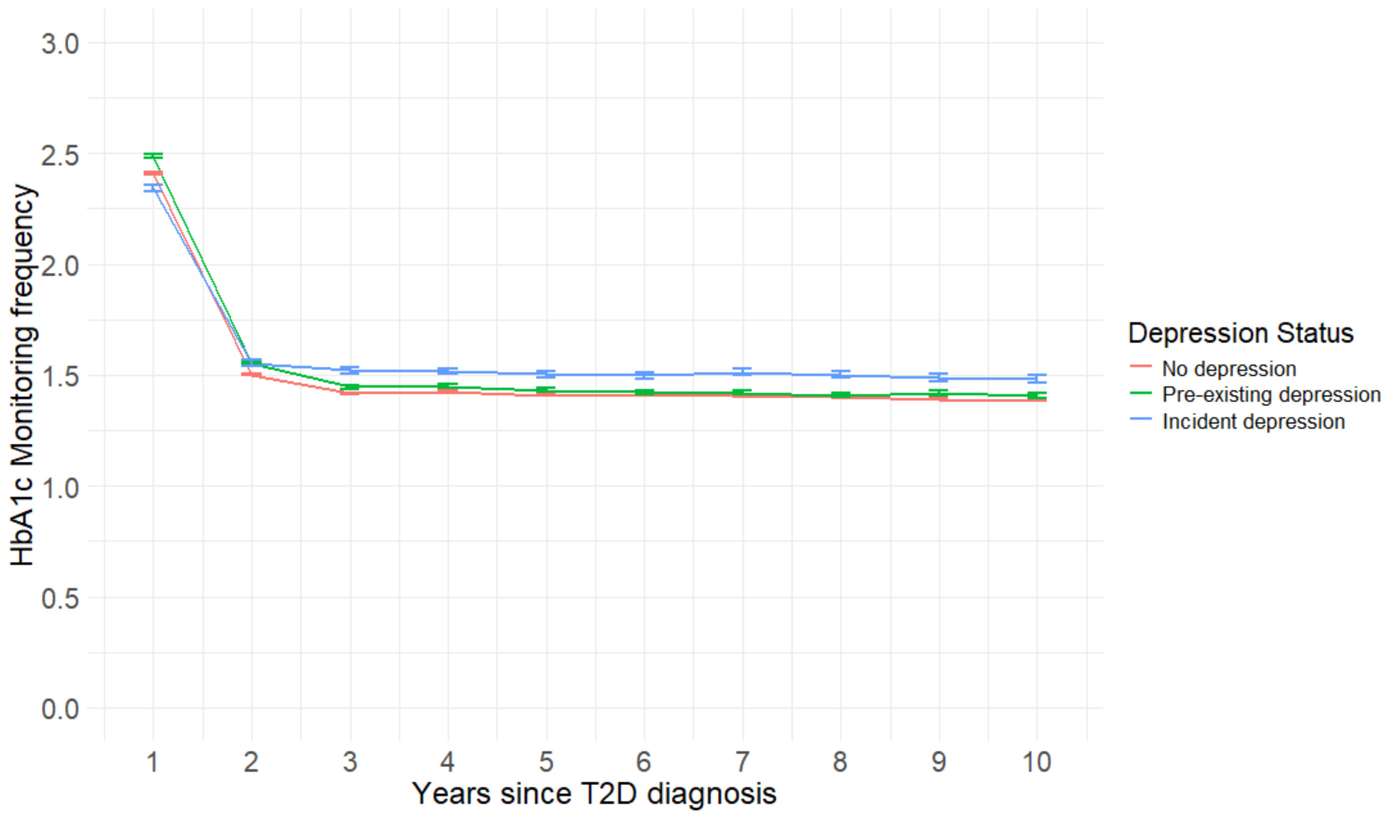

Supplementary figure 6. HbA1c monitoring frequency, stratified by time between T2D diagnosis and depression diagnosis, compared to individuals who do not have a pre-existing depression diagnosis and never receive a diagnosis of incident depression. Error bars represent 95% confidence intervals. Dashed lines represent the time window in which individuals in the incident depression group received a diagnosis. n, number of incident depression cases.

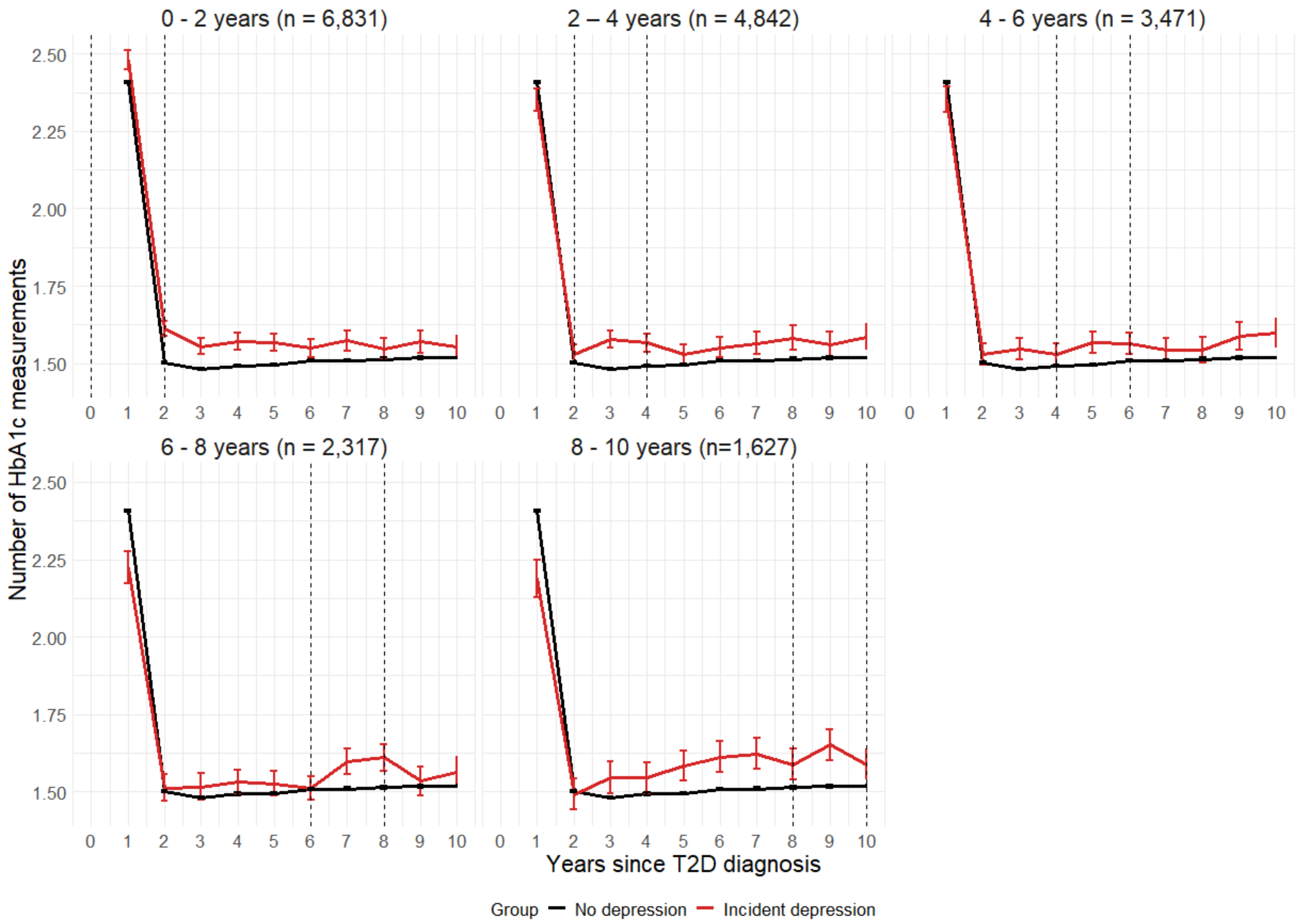

Supplementary figure 7. Time-stratified Cox proportional hazards models for the HbA1c testing frequency categories. Error bars represent 95% confidence intervals. The dashed line represents a hazard ratio of 1.

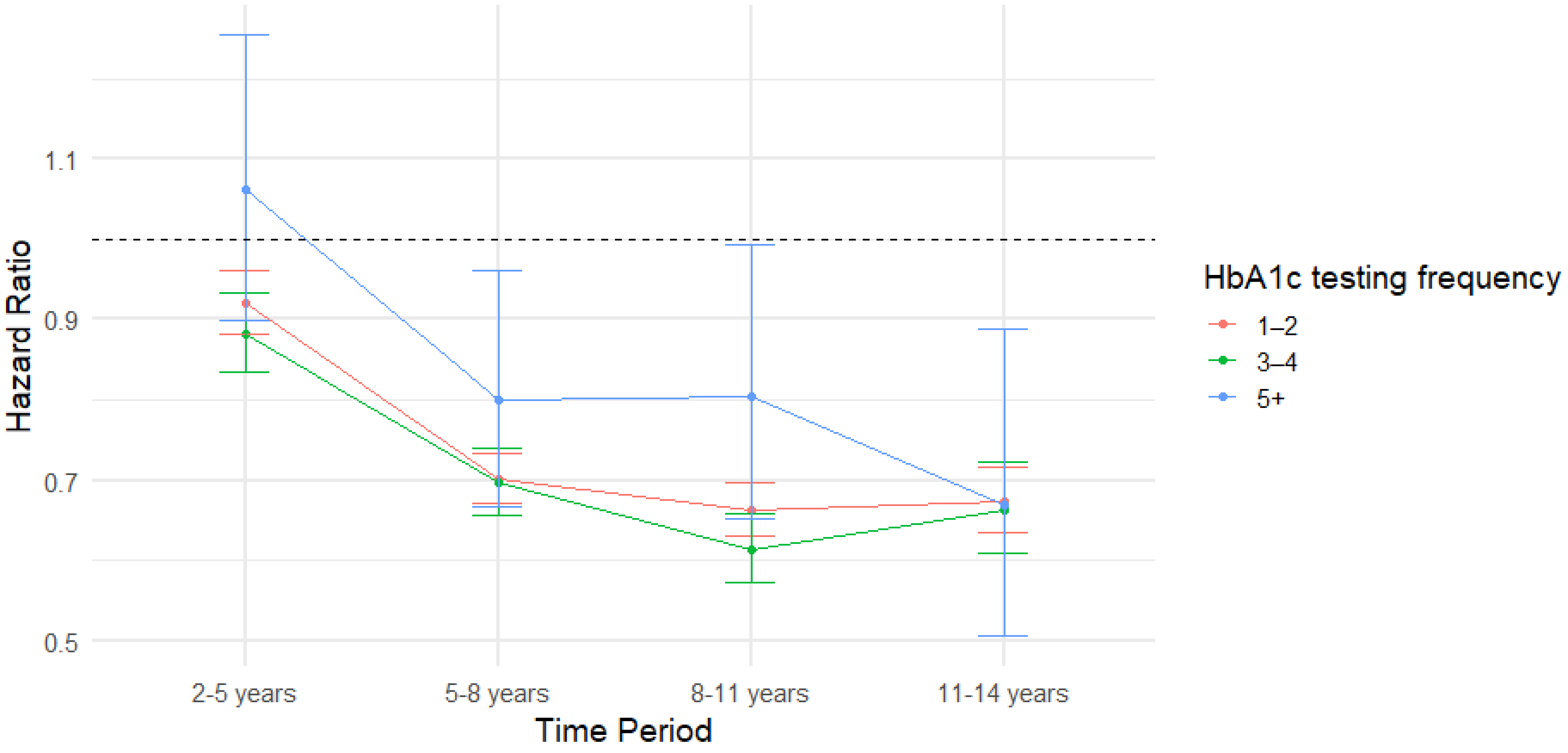

Supplementary figure 8. Time-stratified Cox proportional hazards models for the pre-existing comorbidity score. Error bars represent 95% confidence intervals The dashed line represents a hazard ratio of 1.

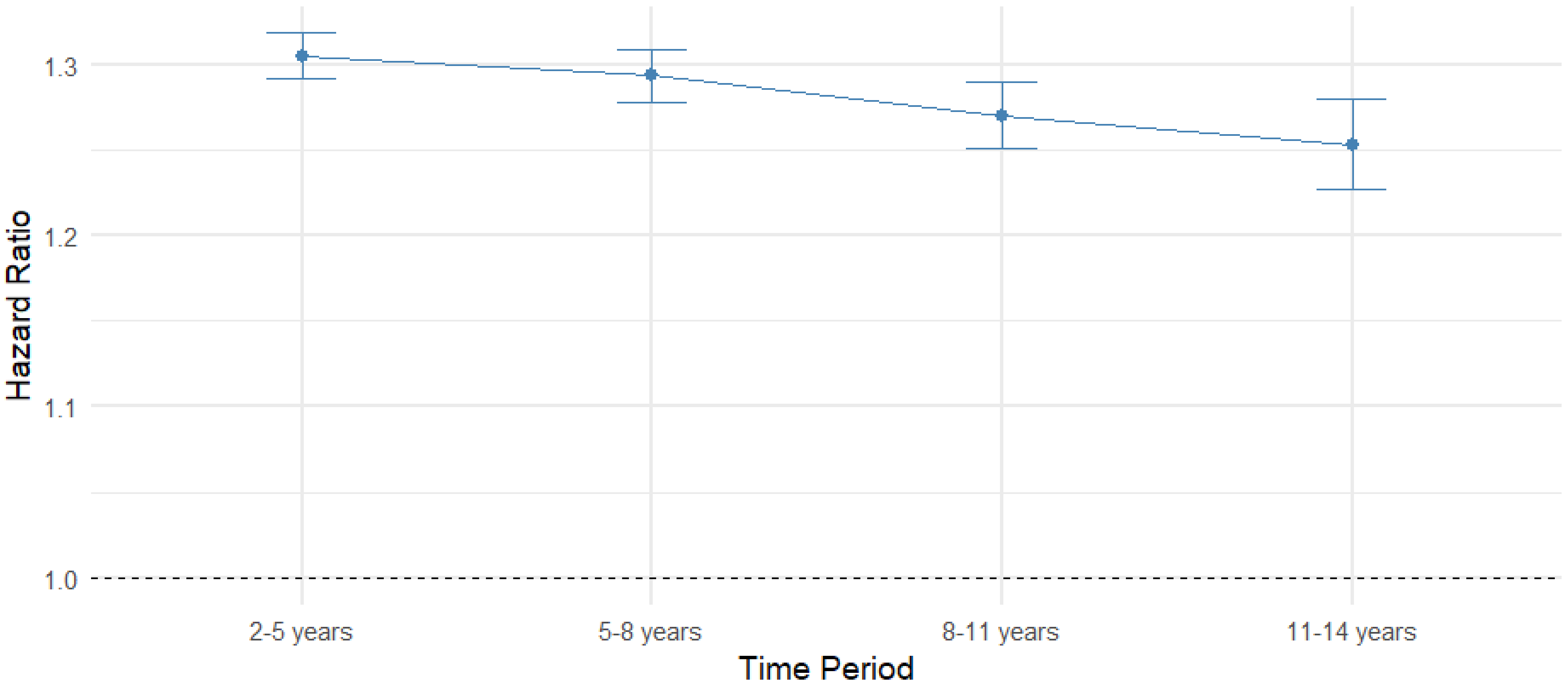

Supplementary figure 9. Time-stratified Cox proportional hazards models for the incident comorbidity score. Error bars represent 95% confidence intervals The dashed line represents a hazard ratio of 1.

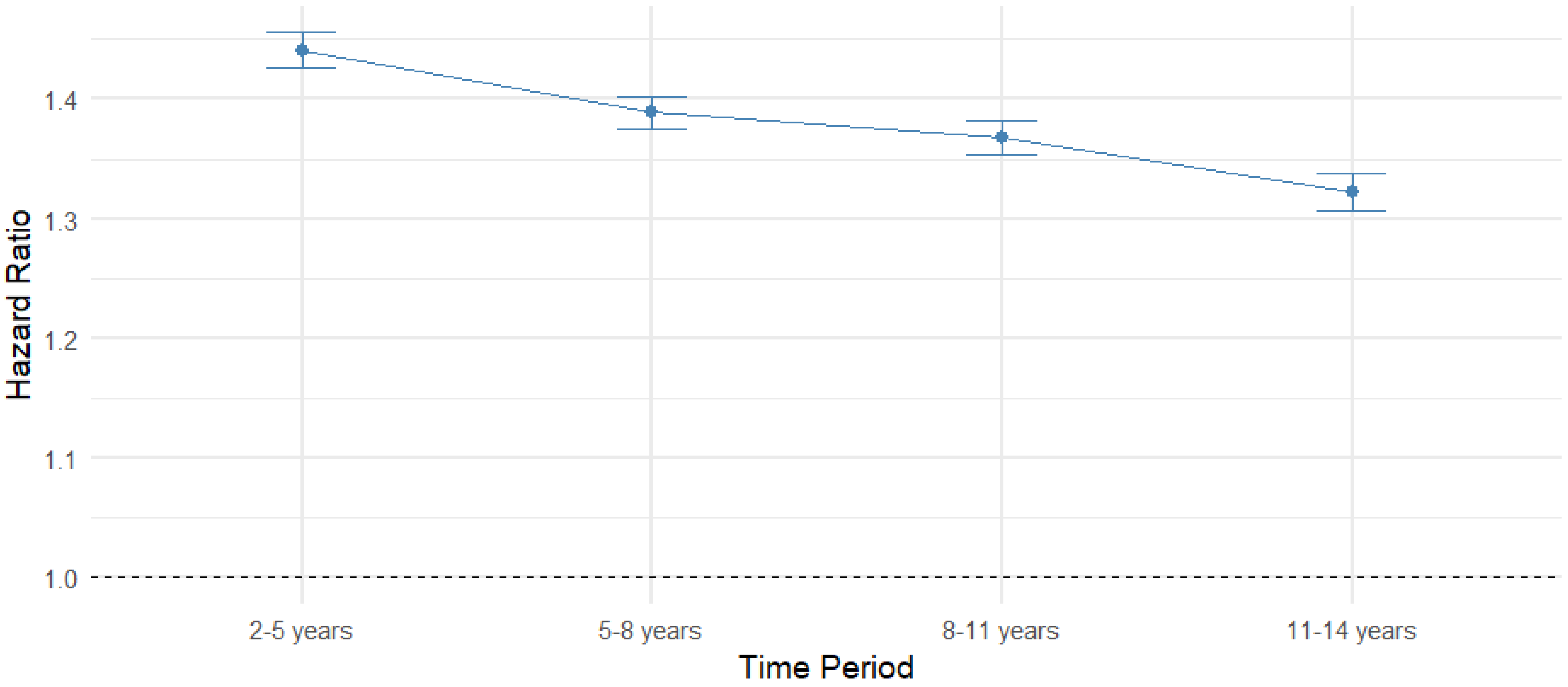

Supplementary figure 10. Time-stratified Cox proportional hazards models for pre-existing depression. Error bars represent 95% confidence intervals. The dashed line represents a hazard ratio of 1.

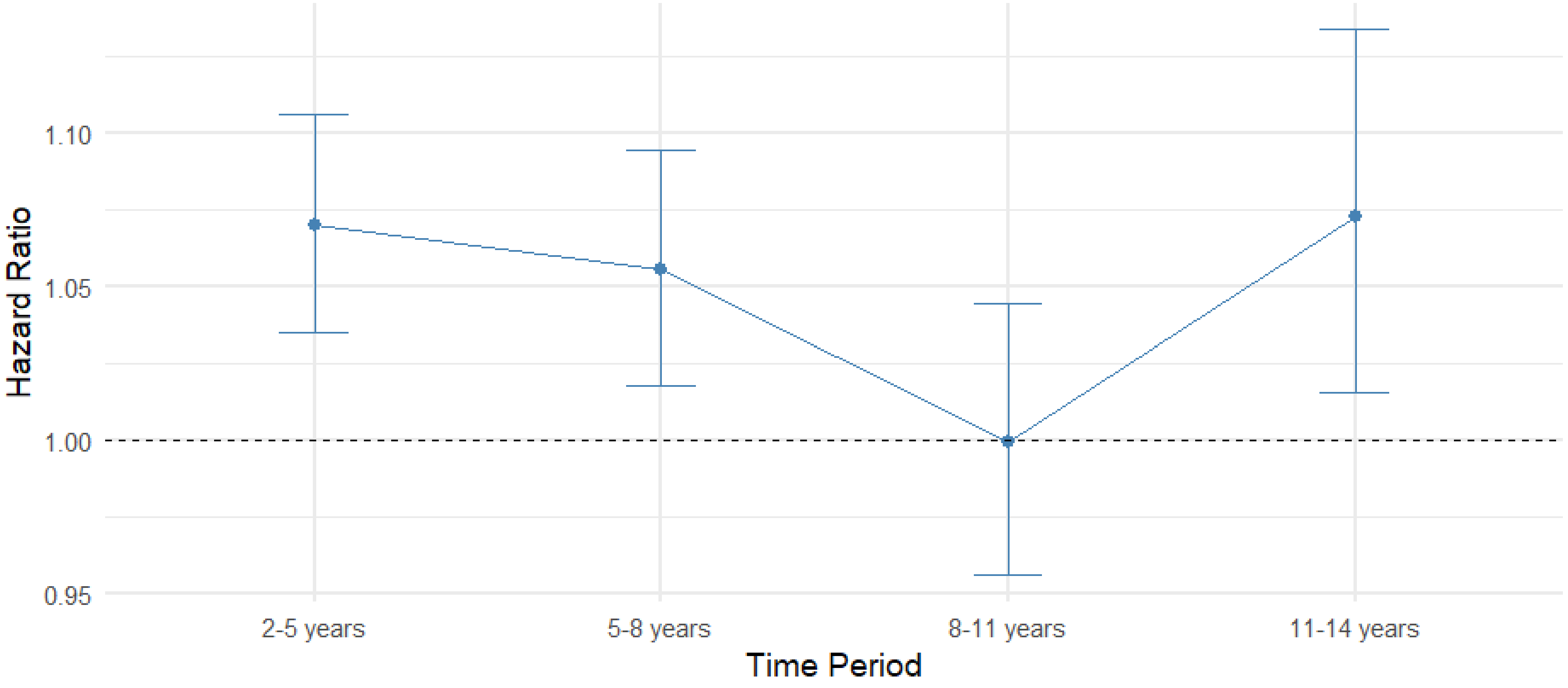

Supplementary figure 11. Time-stratified Cox proportional hazards model for incident depression. Error bars represent 95% confidence intervals. The dashed line represents a hazard ratio of 1.

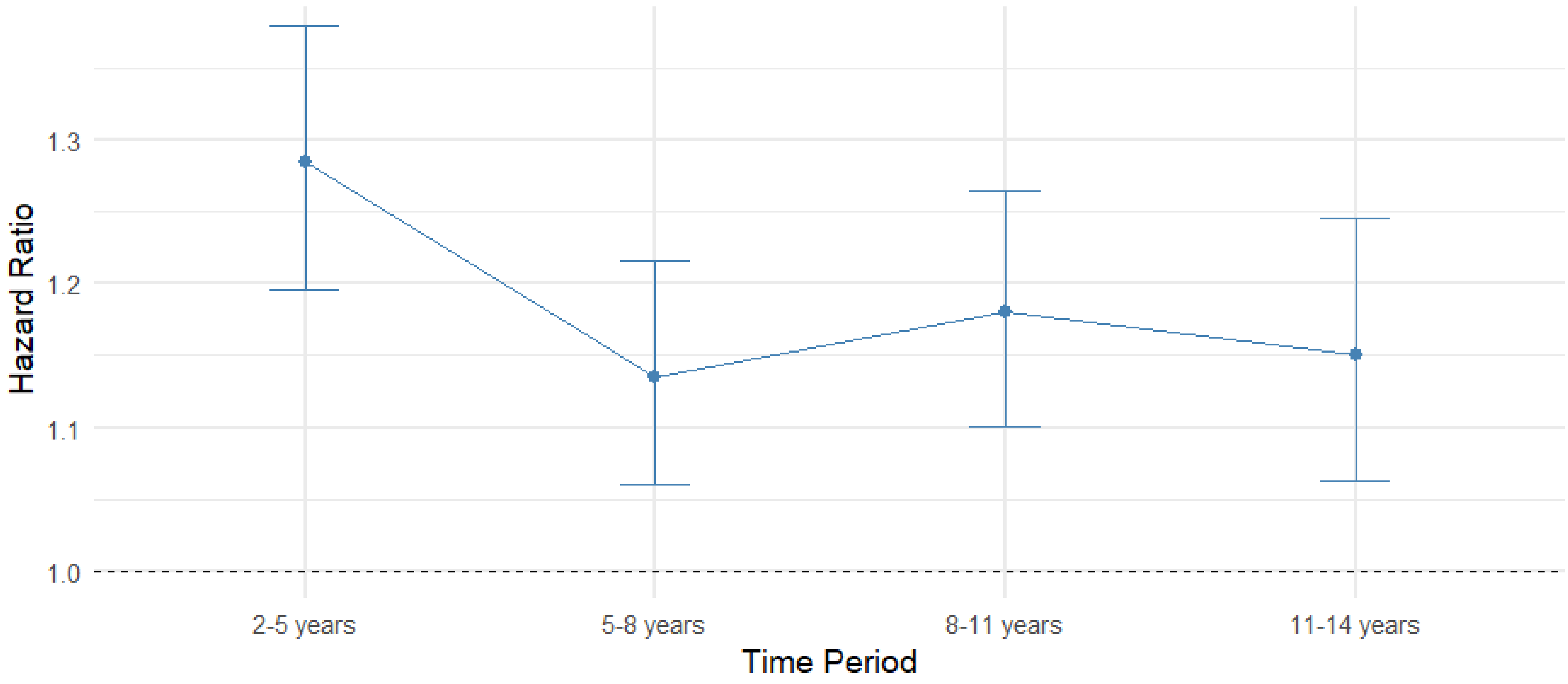

Supplementary Figure 12. The effect of pre-existing depression severity and timing variables on HbA1c monitoring frequency with and without pre-existing and incident comorbidity score adjustment. Error bars represent 95% confidence intervals. *; statistically significant after multiple testing correction ($p < 6.3x10^{-3}$); RD, recurrent depression; TNDC, total number of depression clinical codes; TSLD, time since last depression code from T2D diagnosis; TSDD, time from depression diagnosis to T2D diagnosis; CS, pre-existing and incident comorbidity scores.

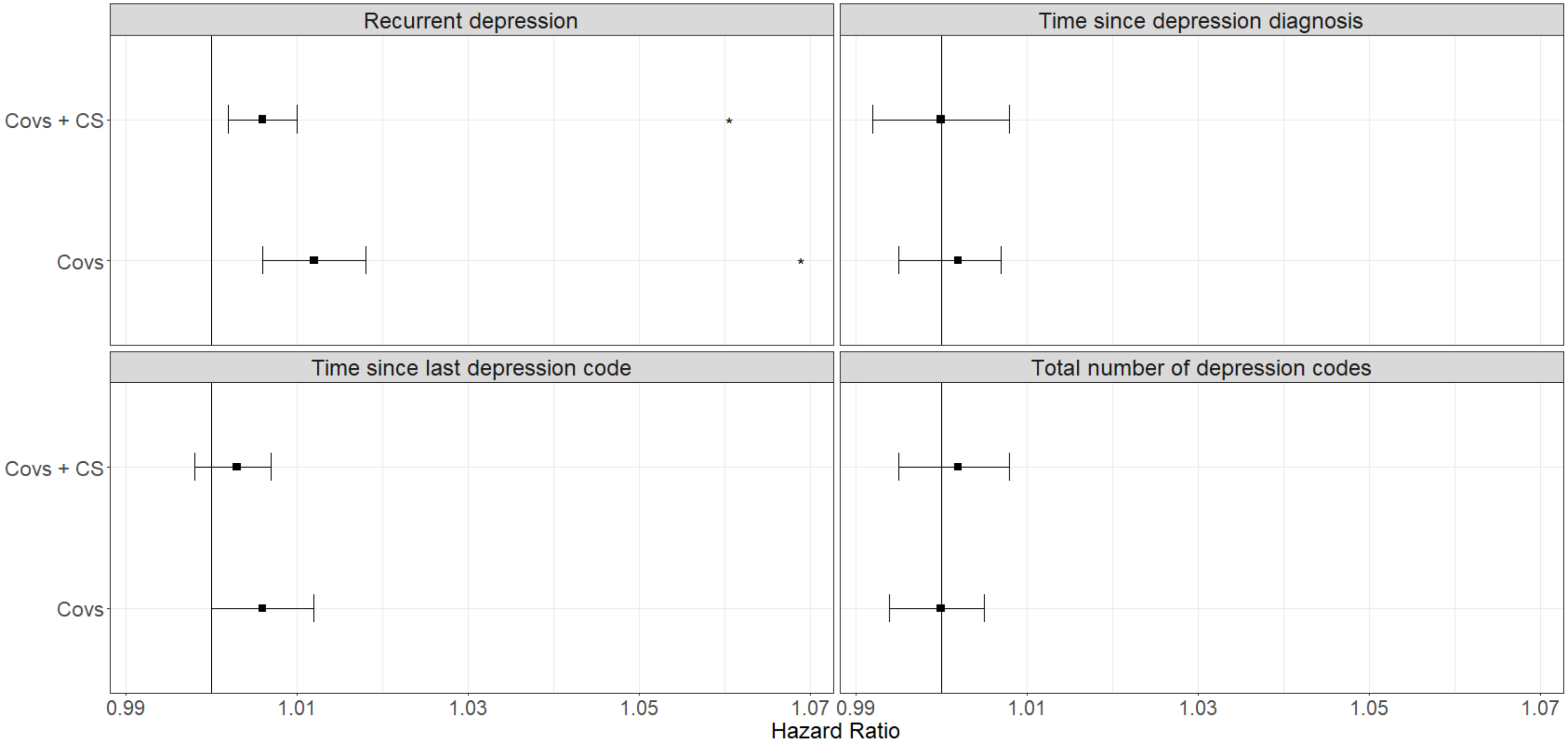

Supplementary Figure 13. The effect of pre-existing depression severity and continuous timing variables on HbA1c monitoring frequency with and without pre-existing and incident comorbidity score adjustment. Error bars represent 95% confidence intervals. Covs, baseline covariates; PreComorbScore, pre-existing comorbidity score; IncComorbScore, incident comorbidity score.*, statistically significant after multiple testing correction ($p < 6.3x10^{-3}$).

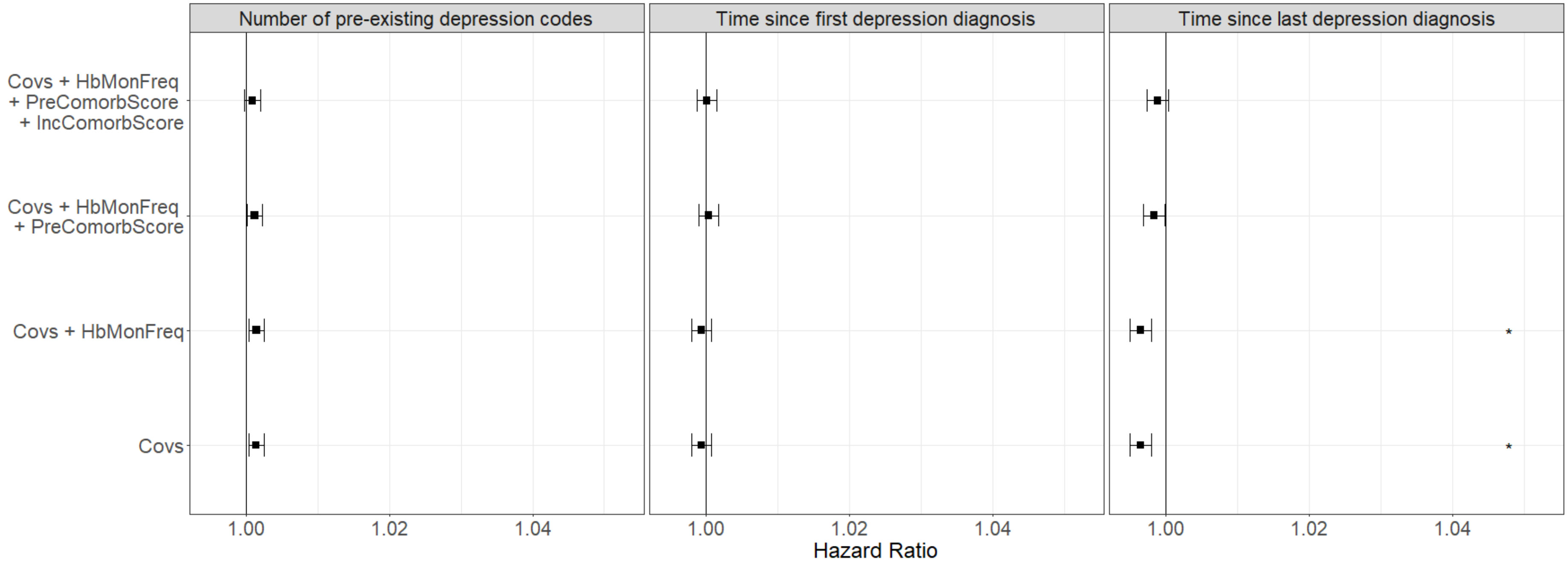

Supplementary Figure 14. The effect of pre-existing depression severity and dichotomous timing variables on HbA1c monitoring frequency with and without pre-existing and incident comorbidity score adjustment. Error bars represent 95% confidence intervals. Covs, baseline covariates; PreComorbScore, pre-existing comorbidity score; IncComorbScore, incident comorbidity score.*, statistically significant after multiple testing correction ($p < 6.3 \times 10^{-3}$).

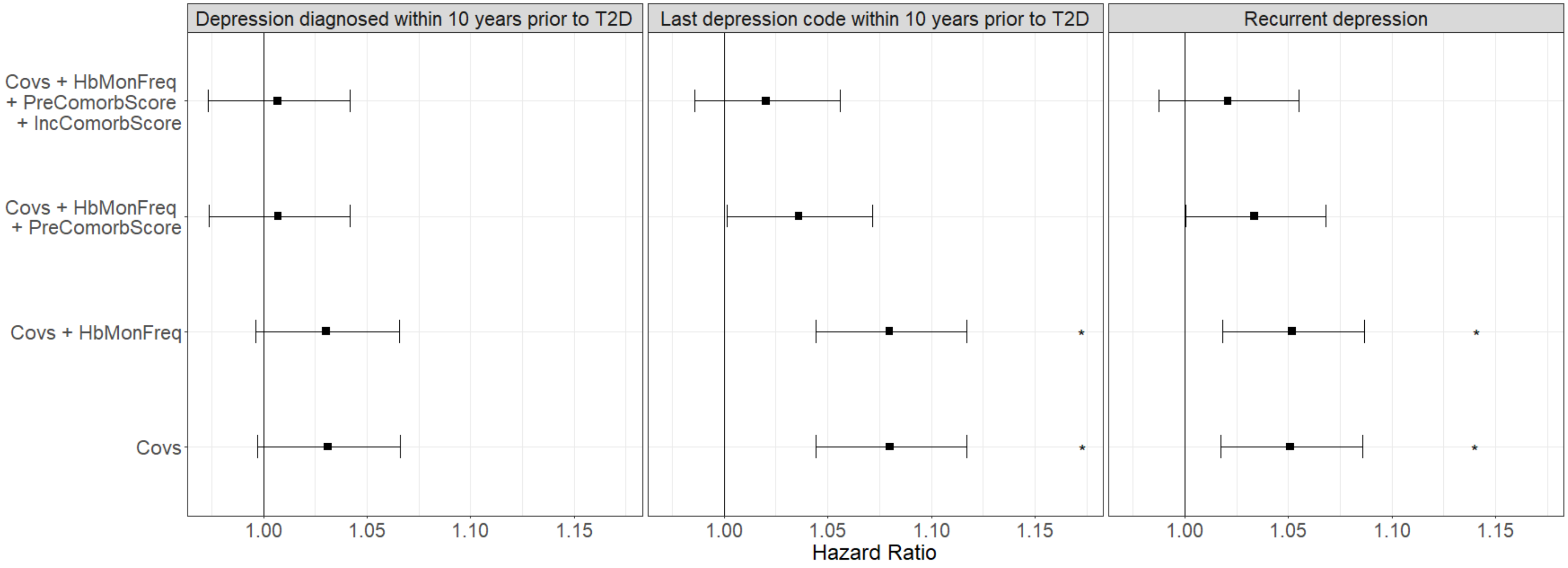

Supplementary figure 15. Primary care healthcare use by year from T2D diagnosis, stratified by depression status. Incident depression represents ever receiving a diagnosis of incident depression during follow up. Error bars represent 95% confidence intervals.

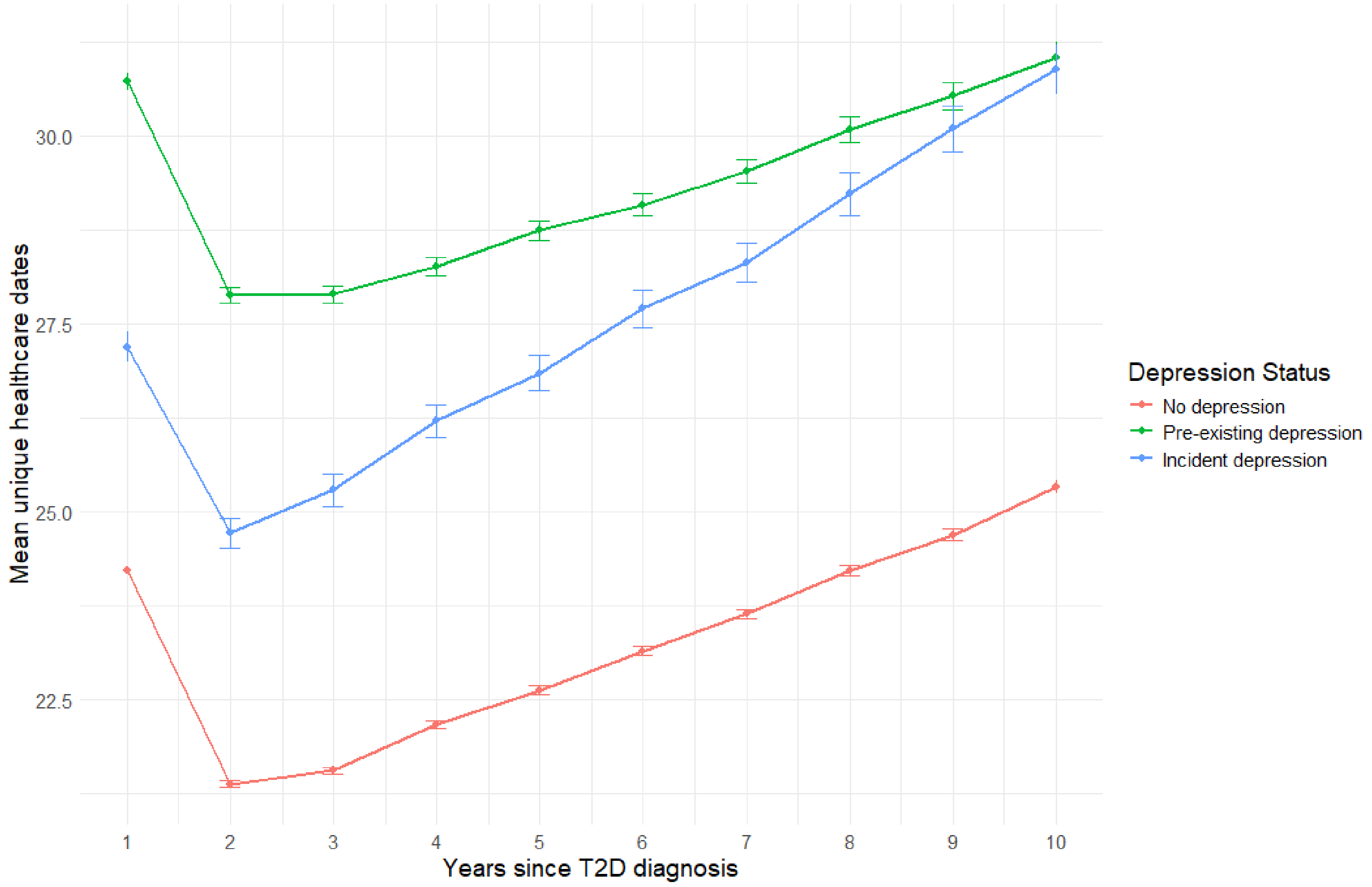

Supplementary figure 16. Primary care healthcare use, stratified by time between T2D diagnosis and depression diagnosis, compared to individuals who do not have a pre-existing depression diagnosis and never receive a diagnosis of incident depression. Error bars represent 95% confidence intervals. Dashed lines represent the time window in which individuals in the incident depression group received a diagnosis. n, number of incident depression cases.

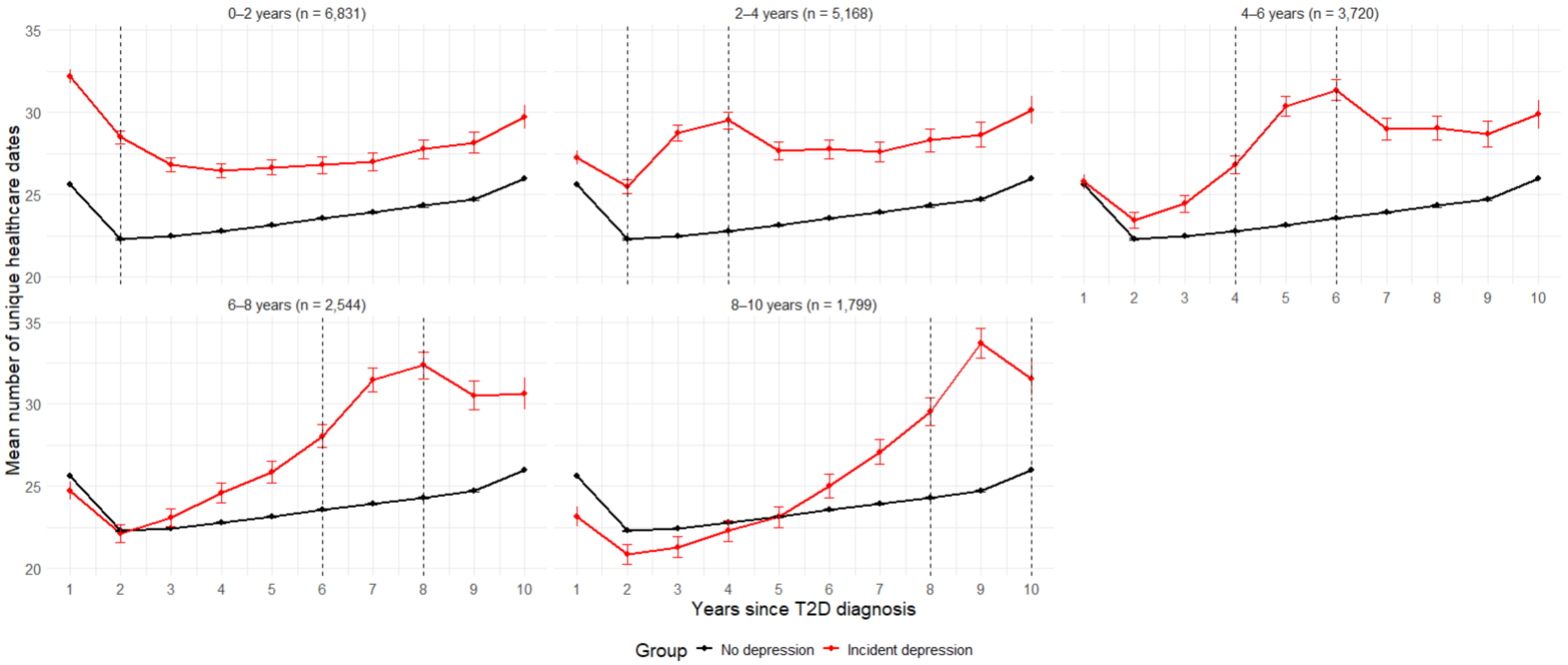

Supplementary figure 17. The association between HbA1c frequency testing categories and risk of all-cause mortality, unadjusted and adjusted for primary care healthcare use. Error bars represent 95% confidence intervals.

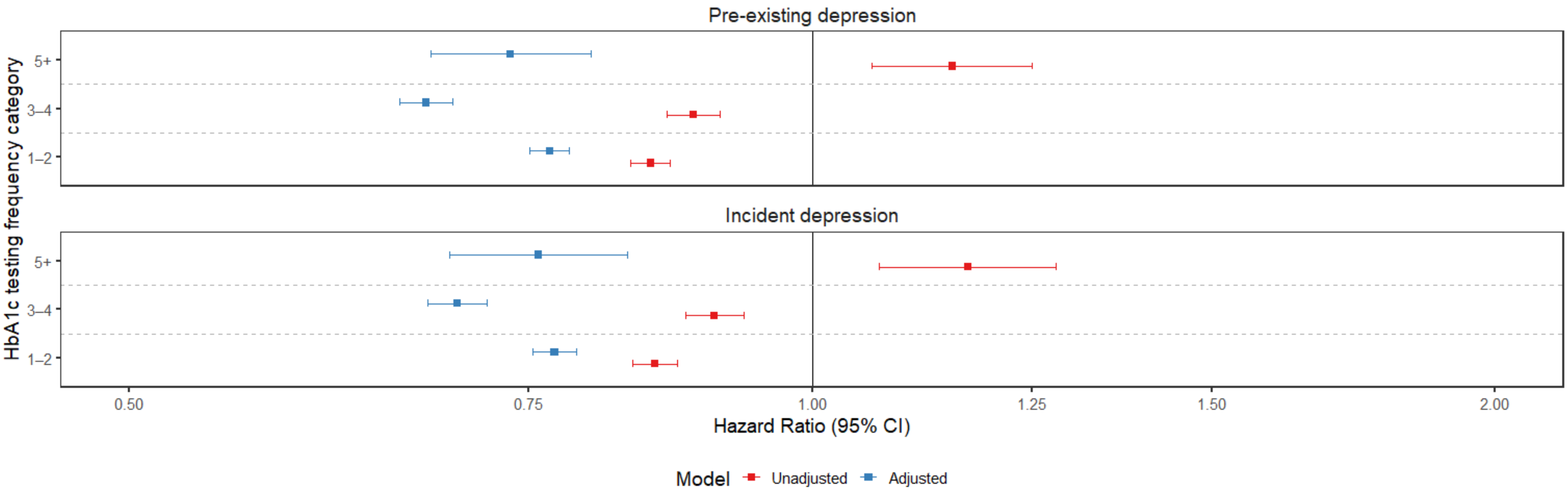
